# Lipoprotein remodelling and elevated serum apolipoprotein B distinguish adults with haemophagocytic lymphohistiocytosis from sepsis

**DOI:** 10.64898/2026.05.13.26352642

**Authors:** Alexandra E Oppong, Keira Louden, Amelia Holloway, Lara Rossi, Thomas McDonnell, George A Robinson, Nishkantha Arulkumaran, Jessica J Manson, Elizabeth C Jury

## Abstract

Haemophagocytic lymphohistiocytosis (HLH) is a rare, life-threatening hyperinflammatory syndrome characterised by uncontrolled immune activation. Patients present with a sepsis-like syndrome, but management requires immune suppression. Reduced high- and low-density lipoprotein cholesterol and hypertriglyceridaemia are reported in HLH, suggesting lipid metabolism disturbances, although in-depth serum metabolomic analysis is lacking. Here, a lipid-focused NMR spectroscopy platform was used to define the serum metabolomic landscape of adults hospitalised with HLH compared to those with sepsis (a key clinical mimic), rheumatic disease (potential HLH drivers/triggers), patients following surgical resection of solid organ cancer (non-infectious acute inflammation controls) and healthy controls (HCs). An HLH-associated metabolomic profile was identified and found to be significantly enriched in lipid metabolism-associated pathways including: ‘Plasma lipoprotein remodelling’, ‘Lipid particles composition’, ‘Glucose homeostasis’ and ‘Amino acid metabolism’. Metabolomic results were validated using matched whole blood RNA-sequencing and the generation of a gene-metabolite interaction network which identified differentially expressed genes enriched in metabolic modules associated with lipid, amino acid, and glucose metabolism, supporting a coordinated metabolic dysregulation in HLH from a transcriptomic to metabolomic level. Compared to all control groups, adults with HLH were defined by elevated apolipoprotein-B (ApoB)-containing triglyceride-rich lipoproteins and saturated fatty acids (area under receiver operator curve >0.70). Validation of a potential HLH-specific lipid signature using ELISA technology identified ApoB (p=0.007) and ApoB:ApoA1 (p=0.0002) as significantly different between HLH and HLH-mimics, including sepsis, supporting their utility as clinically translational biomarkers to distinguish HLH from a key mimic.

## INTRODUCTION

Haemophagocytic lymphohistiocytosis (HLH) is a life-threatening hyperinflammatory syndrome characterised by uncontrolled activation and proliferation of macrophages and CD8^+^ cytotoxic T-cells and impaired cytotoxic cell function^1^. Clinical manifestations include unremitting fever, hepatosplenomegaly, lymphadenopathy, cytopenia, elevated liver enzymes, hyperferritinaemia, hypertriglyceridemia, hypofibrinogenemia and haemophagocytosis^2^. In adults, HLH is usually secondary to an underlying condition such as infection, malignancy, autoimmune/autoinflammatory disease, pregnancy and following treatments such as bone marrow transplant or CAR-T therapy. Defects in cytotoxic lymphocyte function are thought to be acquired and the condition has traditionally been regarded as occurring in the absence of an identifiable genetic defect in cytotoxicity, although recent reports support that primary genetic defects lead to a first episode of HLH in a subset of adults^3^.

Dyslipidaemia can induce changes in immune cell function and has long been linked with inflammation in association with atherosclerosis, where most of the information about the inter-relationship of serum lipid metabolites and immune cell function is reported^4,5^. There is evidence to support that dyslipidaemia can directly influence the homeostasis of plasma membrane lipid rafts, which play an essential role in immune cell function via controlling intracellular signalling events at the immune synapse^6^. Altered lipid metabolism in HLH is under-investigated and while HLH is frequently characterised by hypertriglyceridemia, this feature is not observed consistently across all patients, with several studies reporting variable incidence^7,8^. Disrupted cholesterol metabolism has also been identified, characterised by reduced levels of serum total cholesterol, low density lipoprotein—cholesterol (LDL-C) and high-density lipoprotein-cholesterol (HDL-C)^7–10^. Other lipid abnormalities described in HLH include increased serum secretory sphingomyelinase and bioactive sphingolipids, including elevated C_16_-ceramide:sphingosine ratio^11^. Dyslipidaemia is likewise reported in conditions that can either mimic or trigger HLH. This includes sepsis, which is associated with significantly reduced HDL-C^12^, and chronic inflammatory diseases, such as systemic lupus erythematosus (SLE) which is associated with elevated LDL-C and triglycerides and reduced HDL-C and rheumatoid arthritis (RA) which is associated with decreased LDL-C, and HDL-C^4^.

Our objective was to determine whether a distinct pattern of dyslipidaemia occurs in HLH, which would distinguish HLH from other key differentials^13,14^. We used a lipid-focused metabolomic platform to characterise the serum metabolome of adults hospitalised with HLH and to identify potential metabolomic biomarkers that distinguish HLH from HLH-mimics including sepsis, rheumatological disease triggers (SLE and RA) and following surgical resection of solid organ cancer (as a non-infectious acute inflammation control).

## METHODS

### Patient and control cohorts (Table 1, Figure 1 for study overview)

All patients were recruited through University College London Hospitals NHS Trust (UCLH).

**Figure 1:**
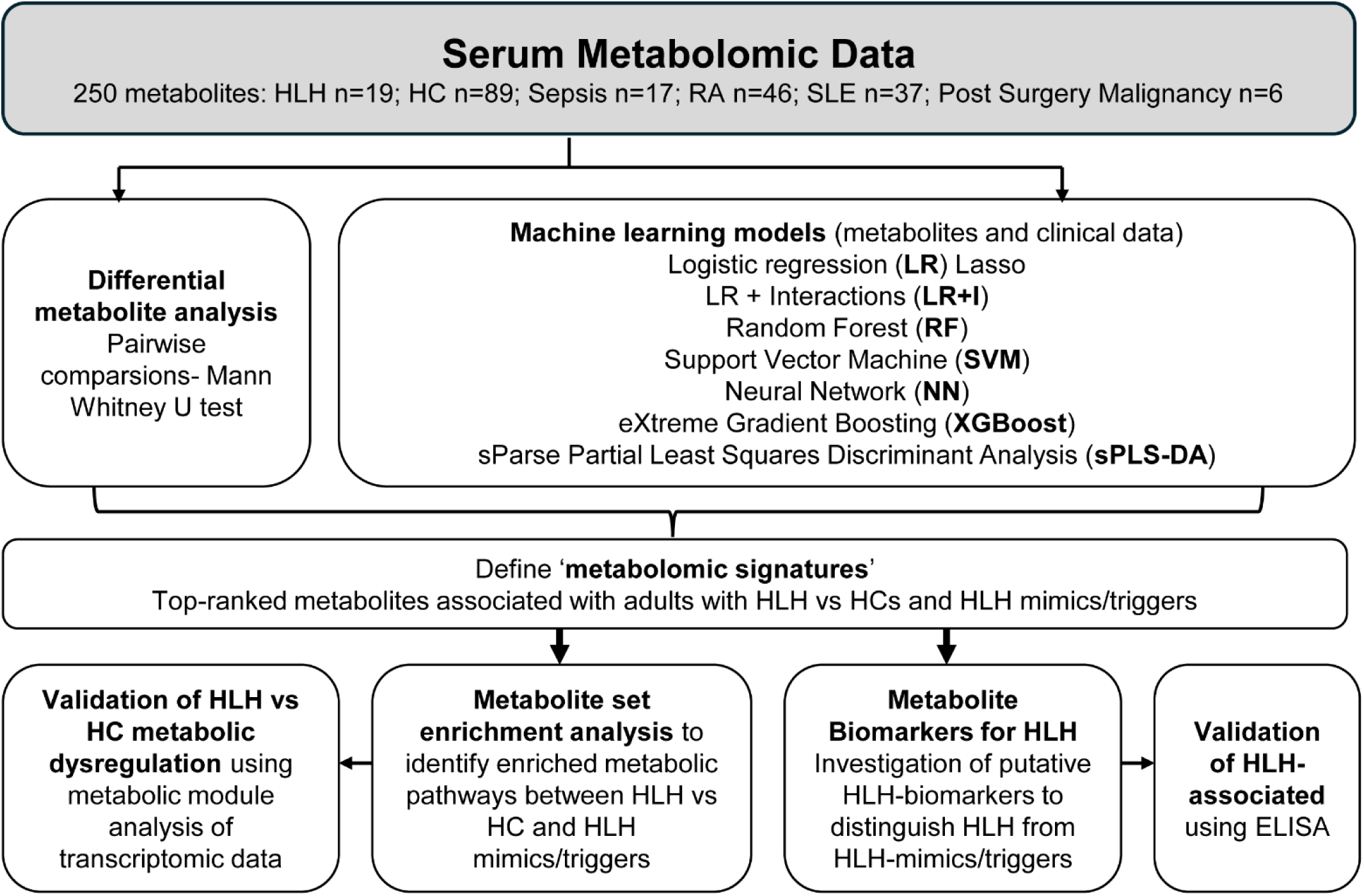
Study Overview. Data analysis pipeline for metabolomic data. Analysis was performed on adults hospitalised with HLH. Metabolomic analysis utilised supervised machine learning models and statistical approaches to identify signatures associated with HLH, compared with healthy controls (HCs), and disease controls (sepsis, systemic lupus erythematosus [SLE], rheumatoid arthritis [RA], and post-surgery for malignancy [PS-M]). Metabolomic signatures associated with HLH were assessed for metabolic pathway enrichment to give molecular mechanism insight as well as for potential HLH biomarkers.

**Table 1:**
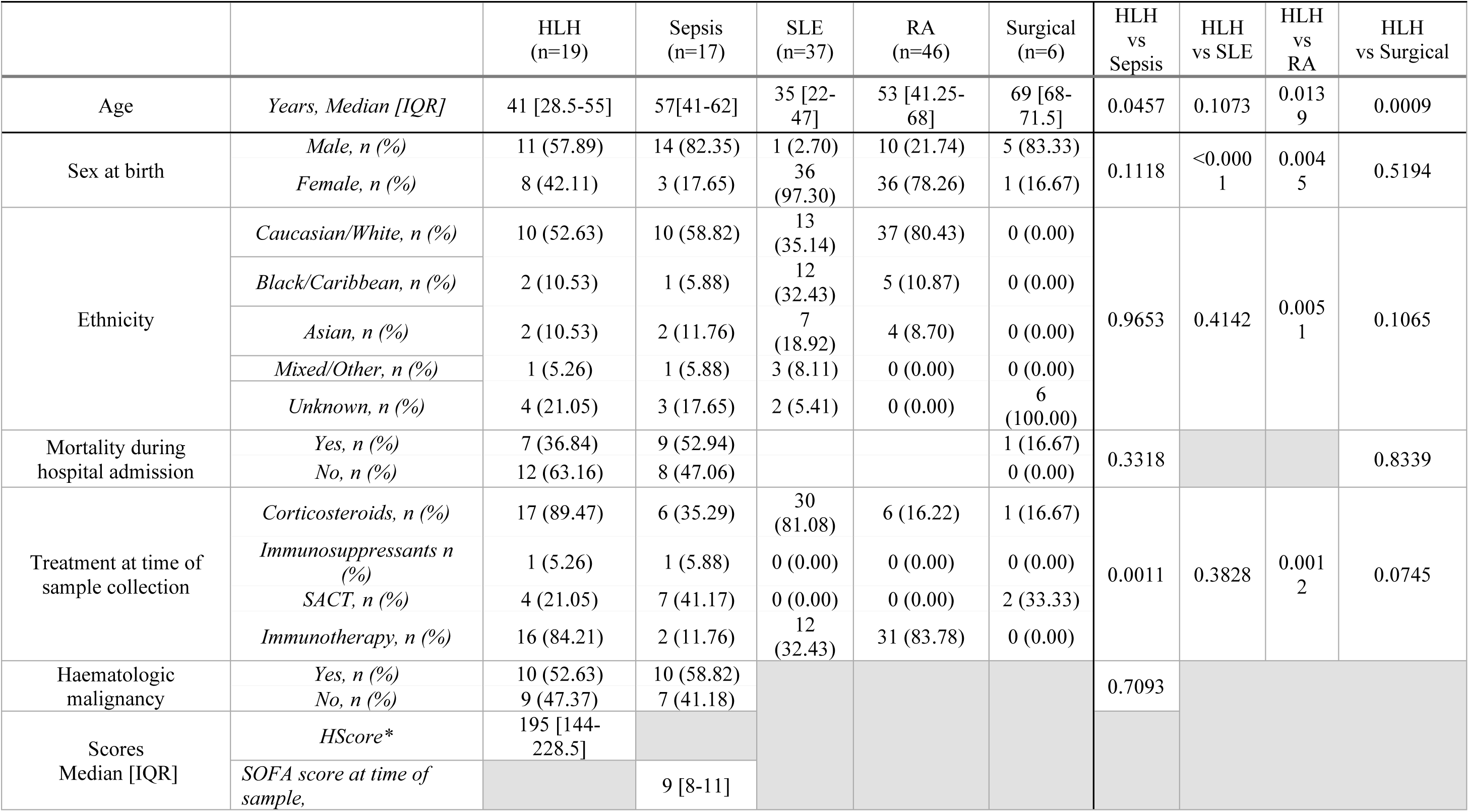
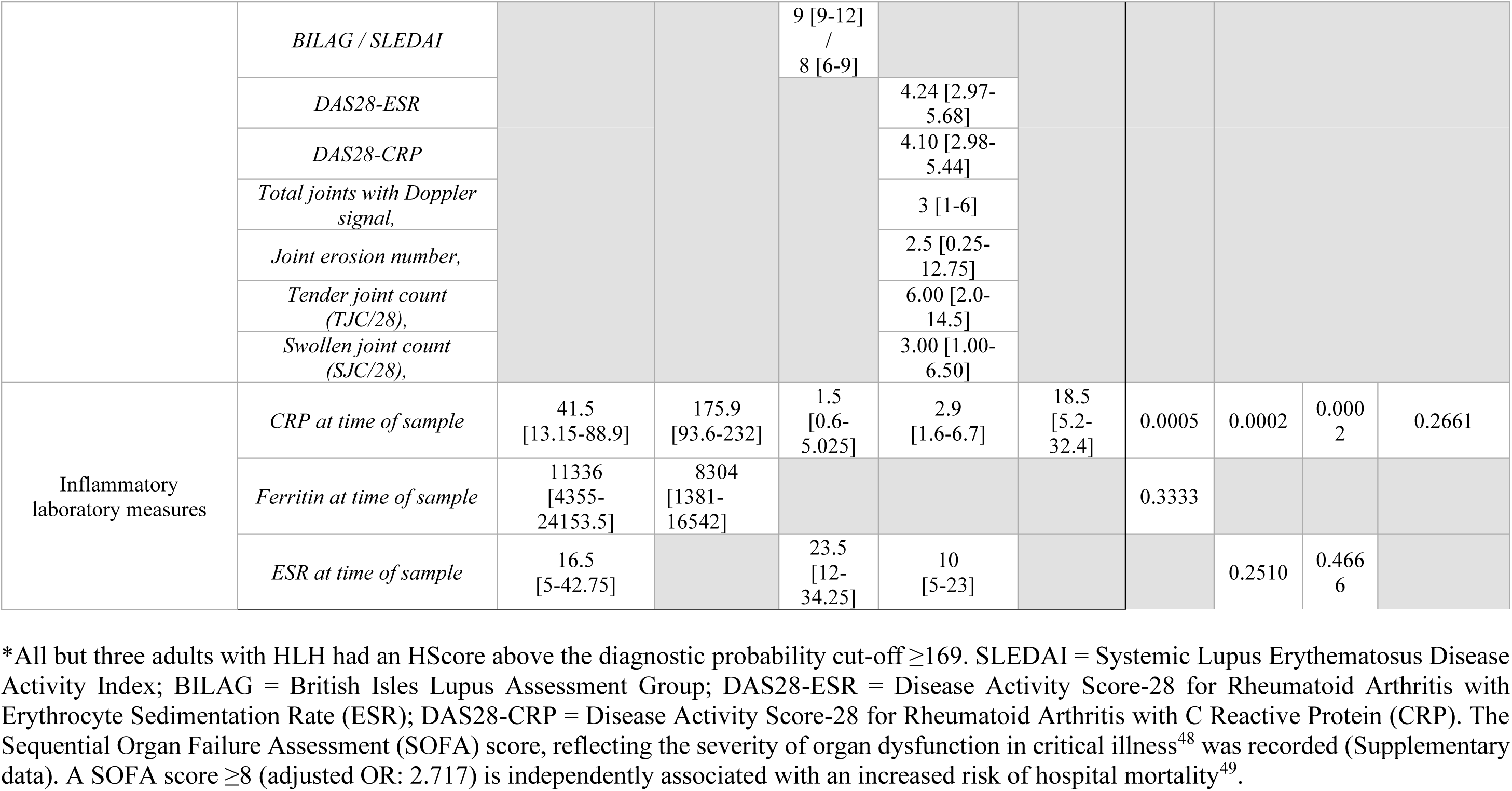
Demographic Table comparing HLH with sepsis, rheumatic and malignancy disease controls.

#### HLH

Serum was collected from adults hospitalised with HLH (n=19) at the tertiary/quaternary UCLH HLH service. Patients were identified using the HScore^15^ and diagnosis confirmed in multidisciplinary team discussions (https://www.uclh.nhs.uk/our-services/find-service/medical-specialties-1/hlh-service). Demographic features and clinical characteristics at the time of sample collection were recorded including HLH trigger, treatment, HScore and routine clinical assay data (Table 1, Table S1).

#### Sepsis and surgery

Patients admitted to the intensive care unit (ICU) at UCLH with suspected or proven bacterial infection and organ dysfunction (n=17) were included. Serum was obtained at time of blood cultures on ICU admission. Serum was also obtained from patients one day following major elective surgical resection of solid organ cancer (n=6).

See Supplemental Methods for details of healthy control (HC), SLE and RA cohort.

#### Ethical Approval

All participants were recruited after gaining informed written consent. Adults with HLH were recruited with approval from Biobank Ethical Review Committee UCLH (15/YH/0311). Patients with sepsis and following surgery were recruited under Research Ethics Committee reference 20/LO/1024.

### Serum metabolomics

Measures of 250 serum biomarkers were acquired with an established nuclear magnetic resonance (NMR)-spectroscopy platform (Nightingale Health, 350μL platform) (Supplemental Methods and Table S2). Metabolites included absolute concentrations, percentages, and ratios of lipoprotein composition. Serum lipids included apolipoproteins (Apo), (very) low-density ((V)LDL), intermediate-density (IDL) and high-density (HDL) lipoprotein particles of different sizes from chylomicrons and extremely large (XXL), very large (XL), large (L), medium (M), small (S), to very small (XS)^16^.

### Statistical analyses

Statistical comparisons of demographic, clinical and treatment data were by Mann-Whitney, Chi-squared or Fisher’s exact test as appropriate.

#### Pairwise comparisons

between (i) HLH vs HCs, and (ii) HLH vs disease control groups (sepsis, RA, SLE and post-surgery malignancy) were by multiple Mann-Whitney U tests. Correction for multiple comparisons was by Benjamini-Hochberg false discovery rate (FDR) (adjusted p-values, padj<0.05 unless stated). Volcano plots visualised differential metabolite abundance between groups, plotting log2 fold change (log2FC), calculated using the median for each metabolite/group, versus p-values (Mann-Whitney U tests).

### Data analysis (Supplemental methods)

#### Predictive models

To derive a combination of serum metabolites/clinical features discriminating adults with HLH from controls, six machine learning (ML) models were implemented: logistic regression (LR), LR with interactions (LR+I); eXtreme Gradient Boosting (XGBoost); random forest (RF); support vector machine (SVM) and neural network (NN)^16^. Sparse Partial Least Squares Discriminant Analysis (sPLS-DA) was used to validate features identified by other methods.

#### Data Handling

Metabolomic data underwent data imputation/homology reduction prior to ML analysis. Homology-reduced datasets included were: HLH vs. HC (n=73), HLH vs. sepsis (n=92), HLH vs. SLE (n=63), HLH vs. RA (n=78). Cohort information was included where available: age, sex, ethnicity, treatment, sodium, potassium, creatinine, estimated glomerular filtration rate, alanine aminotransferase, albumin, alkaline phosphatase, bilirubin, triglyceride, ferritin, C-reactive protein (CRP), haemoglobin, white cell count, neutrophil, lymphocyte, platelets, red cell count, erythrocyte sedimentation rate (ESR) and international normalised ratio.

### ELISA

Serum levels of ApoB and ApoA1 in adults with HLH and controls were validated using ELISA (Quantakine, Biotechne) (Supplemental methods).

## RESULTS

### Dyslipidaemia in adults hospitalised with HLH is characterised by lipoprotein remodelling

To characterise HLH-associated dyslipidaemia a comprehensive assessment of lipoprotein particle size, lipid composition and other metabolites was undertaken using a serum metabolomic platform (Figure 1). HLH was clearly discriminated from HCs using pairwise comparisons (Figure 2A) and six ML models (applied to combined metabolomic/demographic features) (Table 2, Figure S1A-B). Overall, HLH was associated with depletion of HDL particles and ApoA1; elevation of ApoB-containing lipoprotein particles (LDL, IDL and VLDL); lipoprotein remodelling towards triglyceride enrichment and cholesterol depletion and fatty acid dysregulation compared with HCs (Figure 2B-E). Amino acids, glycolysis-related metabolites, and ketone bodies were also differentially regulated in HLH (Figure S1C-E). The HLH-associated metabolite profile was significantly enriched in multiple pathways related to lipid metabolism including: “Familial hyperlipidaemia type 1-5” (p=1.52E-11), “Lipid particles composition” (p=1.05E-13), “Plasma lipoprotein clearance” (p=2.67E-07), ‘Plasma lipoprotein remodelling” (p=3.96E-06) and “SLC-mediated transmembrane transport” (p=6.74E-12). Other enriched pathways included ‘Glucose homeostasis” (p=4.38E-14) and “Amino acid metabolism” (p=8.99E-10) supporting a wider metabolic disturbance in HLH (Figure 2F).

**Figure 2:**
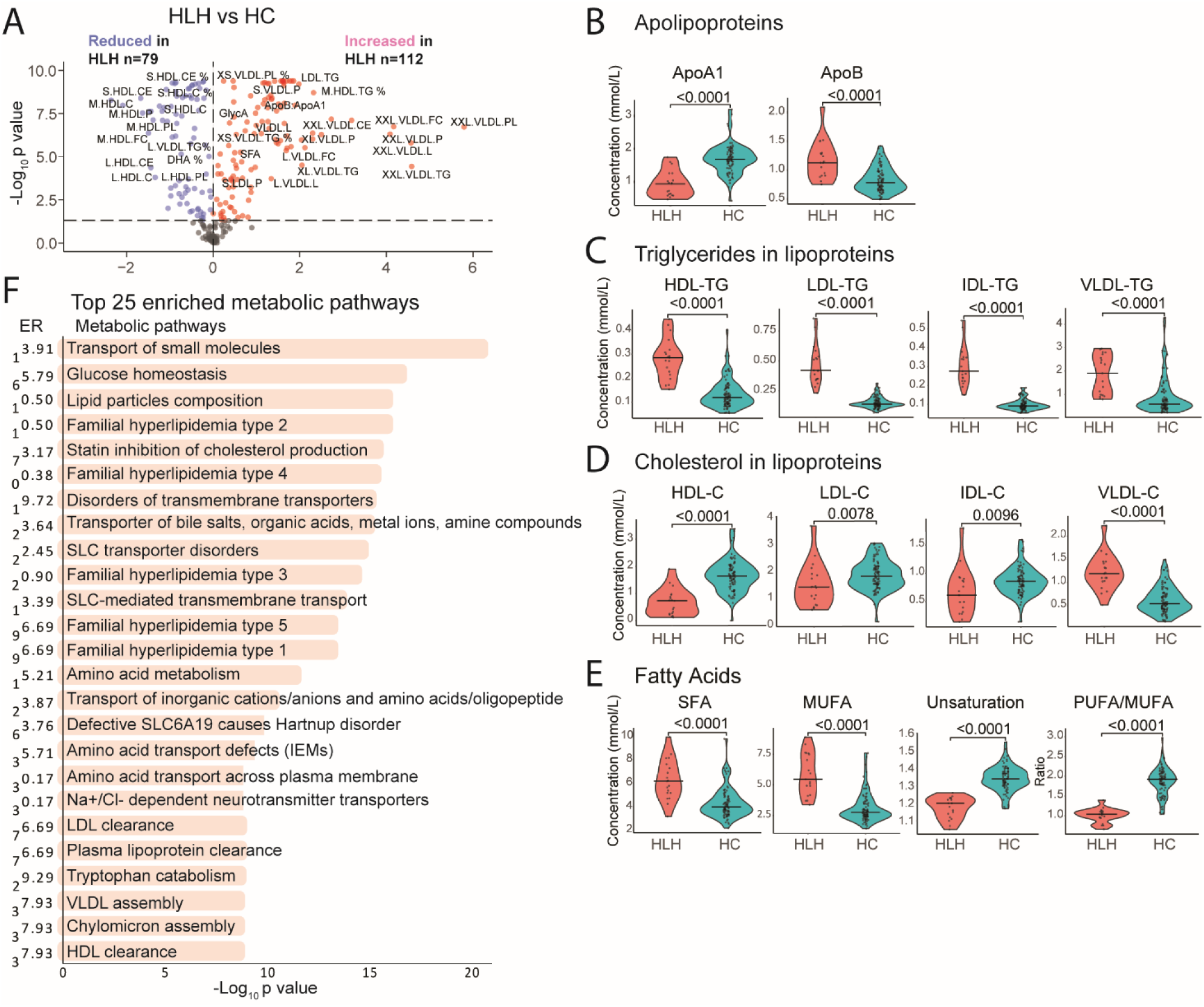
Adults hospitalised with HLH are characterised by changes in lipoprotein composition and remodelling. Sera from adults hospitalised with HLH (n=19) and healthy controls were analysed using an NMR metabolomic platform. (**A**) Metabolomic data were analysed using pairwise comparisons and multiple Mann-Whitney U tests. Volcano plot showing differentially abundant metabolites in HLH vs. HCs; (**B-E**) Violin plots showing the concentration of metabolites in serum from HLH patients (red) and HCs (turquoise) identified by multiple machine learning (ML) models**(B)** apolipoproteins; **(C)** triglycerides in lipoproteins; **(D)** cholesterol in lipoproteins; **(E)** fatty acids. Mann-Whitney U tests, with p-value, each data point and median shown. Refer to Figure S1A-E. (**F**) Metabolites associated with HLH vs HCs identified by ML analysis (≥2 models) and/or differential abundance analysis were analysed by metabolite set enrichment analysis. The top 25 pathways with a significant enrichment (padj< 0.05) are plotted as - log10(raw p-value) on the x-axis. Enrichment ratio (ER) is indicated along the y-axis.

**Table 2:**
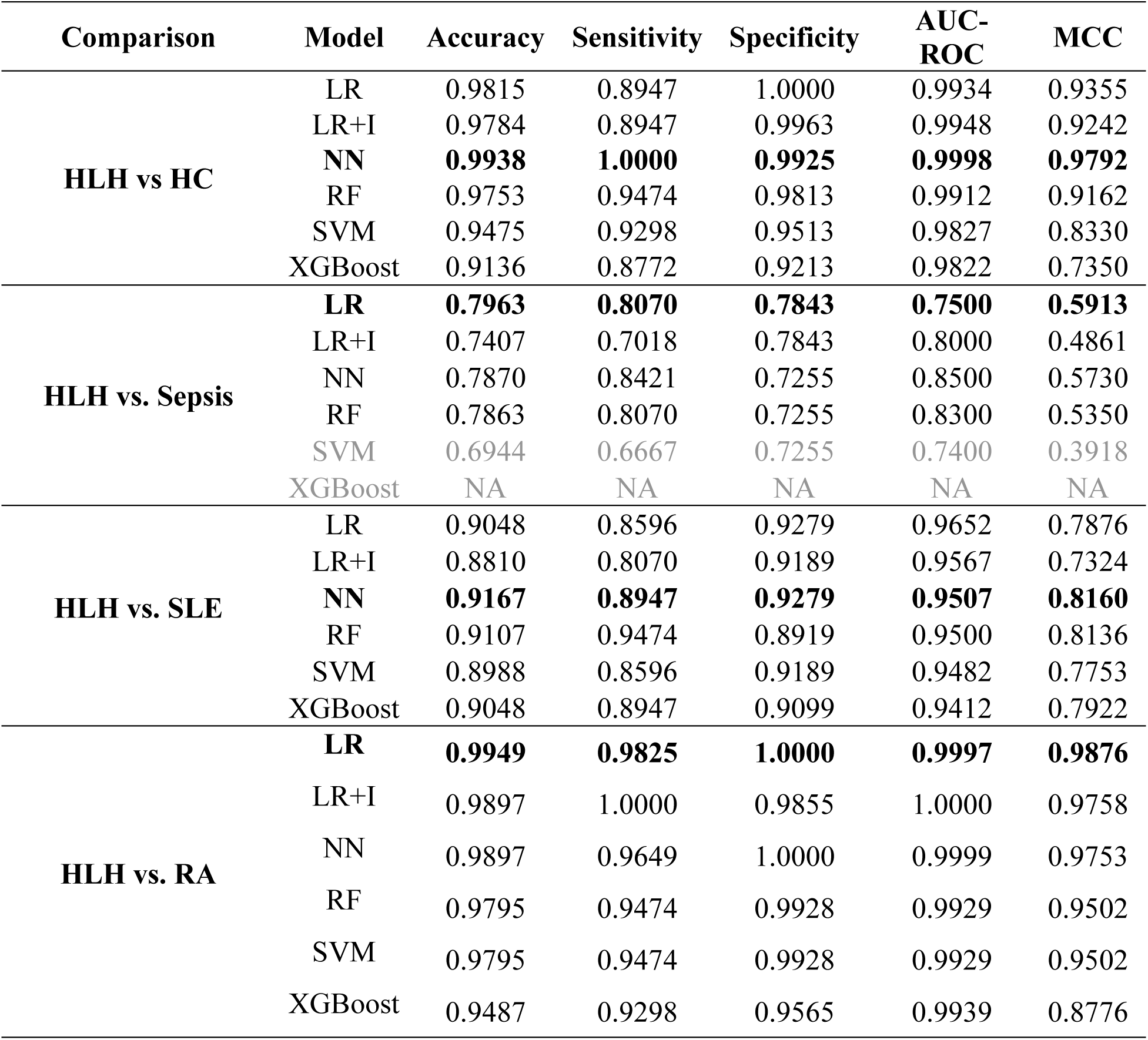
Serum metabolites combined with clinical and demographic classified adults hospitalised with HLH from HC, sepsis (HLH mimic), and other disease controls. Performance statistics for six predictive machine learning models based on serum metabolites and demographic/clinical features from adults with HLH vs Healthy controls (HC, n=89); HLH (n=19) vs. Sepsis (n=17); HLH (n=19) vs SLE (n=37); HLH (n=19) vs RA (n=46). The models used were Lasso logistic regression (LR) with and without interactions (I), neural network (NN), random forest (RF), support vector machine (SVM), and extreme gradient boosting (XGBoost). Statistics were rounded to four decimal places. Greyed out models were excluded from downstream analysis due to poor performance. Top performing models in bold. NA, not available.

| Comparison | Model | Accuracy | Sensitivity | Specificity | AUC-ROC | MCC |
| --- | --- | --- | --- | --- | --- | --- |
| **HLH vs HC** | LR | 0.9815 | 0.8947 | 1.0000 | 0.9934 | 0.9355 |
|  | LR+I | 0.9784 | 0.8947 | 0.9963 | 0.9948 | 0.9242 |
|  | **NN** | **0.9938** | **1.0000** | **0.9925** | **0.9998** | **0.9792** |
|  | RF | 0.9753 | 0.9474 | 0.9813 | 0.9912 | 0.9162 |
|  | SVM | 0.9475 | 0.9298 | 0.9513 | 0.9827 | 0.8330 |
|  | XGBoost | 0.9136 | 0.8772 | 0.9213 | 0.9822 | 0.7350 |
| **HLH vs. Sepsis** | **LR** | **0.7963** | **0.8070** | **0.7843** | **0.7500** | **0.5913** |
|  | LR+I | 0.7407 | 0.7018 | 0.7843 | 0.8000 | 0.4861 |
|  | NN | 0.7870 | 0.8421 | 0.7255 | 0.8500 | 0.5730 |
|  | RF | 0.7863 | 0.8070 | 0.7255 | 0.8300 | 0.5350 |
|  | SVM | 0.6944 | 0.6667 | 0.7255 | 0.7400 | 0.3918 |
|  | XGBoost | NA | NA | NA | NA | NA |
| **HLH vs. SLE** | LR | 0.9048 | 0.8596 | 0.9279 | 0.9652 | 0.7876 |
|  | LR+I | 0.8810 | 0.8070 | 0.9189 | 0.9567 | 0.7324 |
|  | **NN** | **0.9167** | **0.8947** | **0.9279** | **0.9507** | **0.8160** |
|  | RF | 0.9107 | 0.9474 | 0.8919 | 0.9500 | 0.8136 |
|  | SVM | 0.8988 | 0.8596 | 0.9189 | 0.9482 | 0.7753 |
|  | XGBoost | 0.9048 | 0.8947 | 0.9099 | 0.9412 | 0.7922 |
| **HLH vs. RA** | **LR** | **0.9949** | **0.9825** | **1.0000** | **0.9997** | **0.9876** |
|  | LR+I | 0.9897 | 1.0000 | 0.9855 | 1.0000 | 0.9758 |
|  | NN | 0.9897 | 0.9649 | 1.0000 | 0.9999 | 0.9753 |
|  | RF | 0.9795 | 0.9474 | 0.9928 | 0.9929 | 0.9502 |
|  | SVM | 0.9795 | 0.9474 | 0.9928 | 0.9929 | 0.9502 |
|  | XGBoost | 0.9487 | 0.9298 | 0.9565 | 0.9939 | 0.8776 |

These results were validated using whole blood RNA-sequencing and metabolic gene module analysis in a subset of matched blood samples (Supplemental methods). Seventy-five HLH-enriched genes were related to metabolic modules of lipid, amino acid, and glucose metabolism (Figure S1F, Table S3). Gene-metabolite interaction analysis defined an interaction network (comprising 158 up- and 10 down-regulated genes interacting with 17 metabolites), highlighting several metabolic pathways prominent in HLH vs HCs, including cholesterol, fatty acid and lipid metabolism; glycolysis, gluconeogenesis and ketogenesis; amino acid metabolism; iron and glutathione metabolism, haem signalling and the urea cycle (Figure 3A). Lipid metabolism-associated genes were represented within the network including those involved with lipid transport, metabolism, and inflammatory regulation; PUFA-driven inflammation; lipoprotein remodelling, triglyceride turnover, and hormonal control of lipid metabolism; and haem metabolism (Figure 3B-E). Taken together, these findings show that peripheral blood from adults with HLH is characterised by a coordinated metabolic dysregulation in pathways related to lipid, glucose, and amino acid metabolism from a transcriptomic to a metabolomic level (summarised in Figure S2).

**Figure 3:**
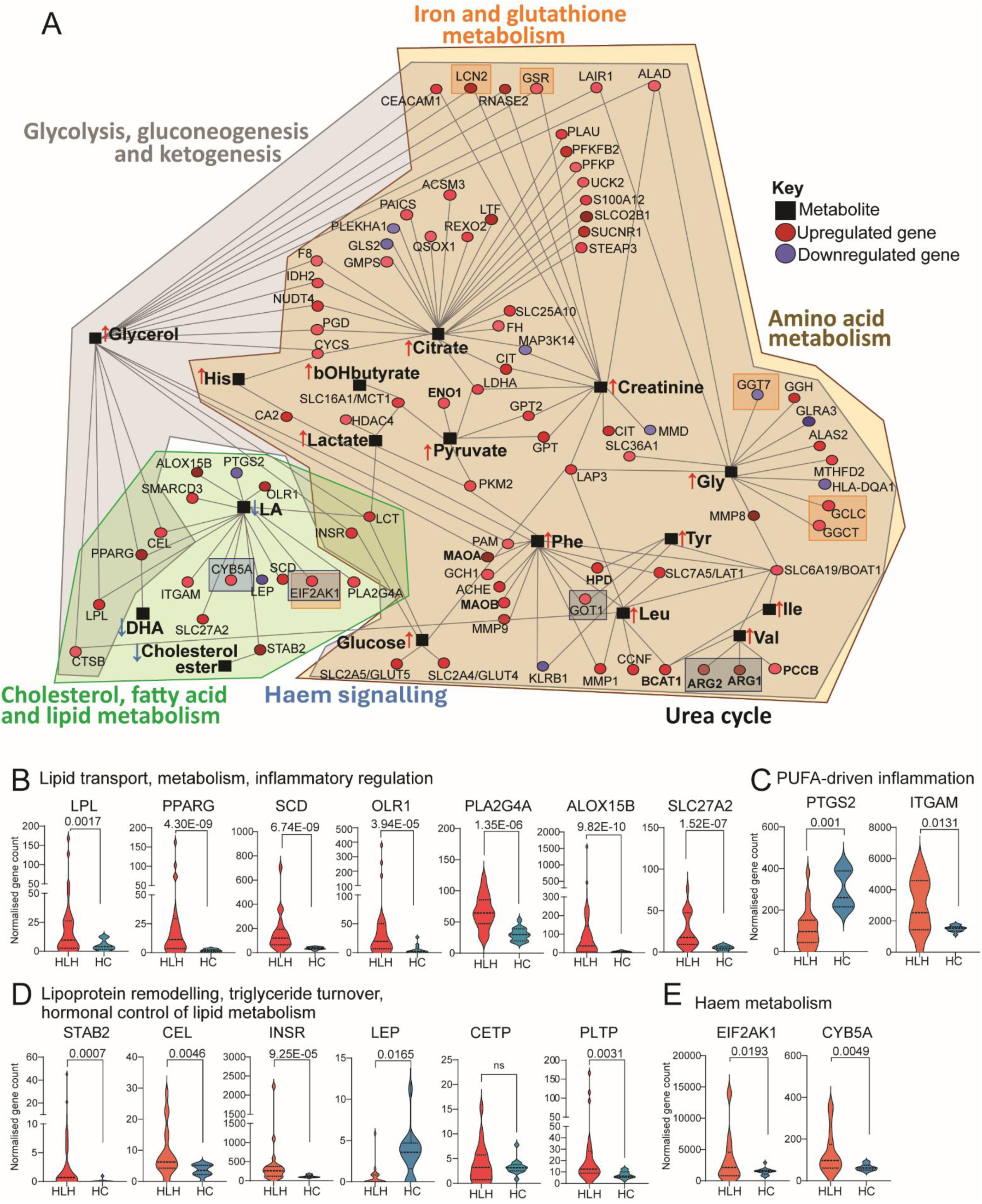
Gene-metabolite interaction network between HLH and HCs. **(A)** Gene-metabolite interaction network illustrating crosstalk between differentially expressed genes (n=158 upregulated, red circles; ten downregulated genes, blue circles) and metabolites (n=17, black squares) in HLH vs HCs using MetaboAnalyst^50^. Associations with metabolic pathways are shown: cholesterol, fatty acid, lipid metabolism (green mask); glycolysis, gluconeogenesis, ketogenesis (grey mask); amino acid metabolism (yellow mask); iron and glutathione metabolism (orange boxes), haem signalling (blue boxes); urea cycle (black boxes). **(B-E)** Violin plots showing genes associated with **(B)** lipid transport, metabolism, and inflammatory regulation (LPL, lipoprotein lipase; PPARG, peroxisome proliferator-activated receptor gamma; SCD, stearoyl-CoA desaturase; OLR1, oxidised low-density lipoprotein receptor 1; PLA2G4A, phospholipase A2 group IVA; ALOX15B, arachidonate 15-lipoxygenase type B; SLC27A2, solute carrier family-27 member-A2); **(C)** PUFA-driven inflammation (PTGS2, prostaglandin-endoperoxide synthase-2; ITGAM, integrin subunit alpha M); **(D)** lipoprotein remodelling, triglyceride turnover, and hormonal control of lipid metabolism (STAB2, stabilin-2; CEL, carboxyl ester lipase; INSR, insulin receptor; LEP, leptin; *CEPT, cholesterol ester transfer protein; *PLTP, Phospholipid transfer protein); **(E)** haem metabolism (EIF2AK1, eukaryotic translation initiation factor-2 alpha kinase-1; CYB5A cytochrome b5 type A). Median, IQR, padj<0.05. *not present in network but associated with lipid metabolism. Refer to Table S3.

### Serum metabolomic signatures and routine patient data can distinguish adults with HLH from those with severe sepsis, autoimmune rheumatic disease, and sterile inflammation

Differentiating HLH from HLH-mimics is key for clinical decision-making, therefore we assessed metabolic differences between adults hospitalised with HLH and those with sepsis. Patients with sepsis were older but had no statistically significant differences in sex, ethnicity or survival (Table 1). Sixty-one metabolites were upregulated and four were downregulated in HLH compared to sepsis (Figure 4A). Of note, levels of VLDL/LDL and triglyceride-rich lipoproteins were elevated, and glycolysis-related metabolites (glucose, pyruvate) were lower in HLH compared to sepsis (albeit higher in HLH and sepsis compared to healthy controls). Four ML models applied to the combined metabolomic, demographic and available routine patient data discriminated well between HLH and sepsis (>70% accuracy) (Table 2, Figure 4B) and outperformed models using metabolites or clinical features alone (Table S4). Overall, 30 metabolite, lipid and routine patient clinical measures were featured in two models or more (Figure 4C). The top features included phospholipids in IDL (IDL-PL, AUC=0.771), glycine (Gly, AUC=0.797), glucose (AUC=0.743), free cholesterol in small HDL (S-HDL-FC, AUC=0.819), corticosteroids (AUC=0.780), triglyceride in large HDL (L-HDL-TG, AUC=0.731), triglyceride in large LDL (L-LDL-TG, AUC=0.732), neutrophil count (AUC=0.724), acetone (AUC=0.762), pyruvate (AUC=0.718), VLDL size (AUC=0.554), M-LDL-CE (AUC= 0.783) and CRP (AUC=0.828) (Figure 4D). Interestingly, the predictive performance of the models was similar when steroid treatment (a classifying feature identified by three ML models) was excluded from the analysis except for the addition of unsaturation (AUC=0.416), acetate (AUC=0.511), and clinically measured creatinine (AUC=0.663) (Table S4, Figure S3A-B). The potential confounding effects of corticosteroid treatment on metabolic profiles^17^ were also assessed by univariate logistic regression. After adjusting for steroid use IDL-PL (Odds ratio [OR]: 4.815, p=0.0177), S-HDL-FC (OR: 4.378, p=0.0198), L-HDL-TG (OR: 3.043, p=0.0341), glucose (OR: 0.264, p=0.0382), pyruvate (OR: 0.4314, p=0.0662), and acetone (OR:25.93, p=0.0540) were still significantly altered between HLH and sepsis. Together, the top 12 metabolomic and clinical features effectively clustered HLH from sepsis showing that metabolic markers could discriminate between patients with different hyperinflammatory conditions during active disease (Figure 4E).

**Figure 4:**
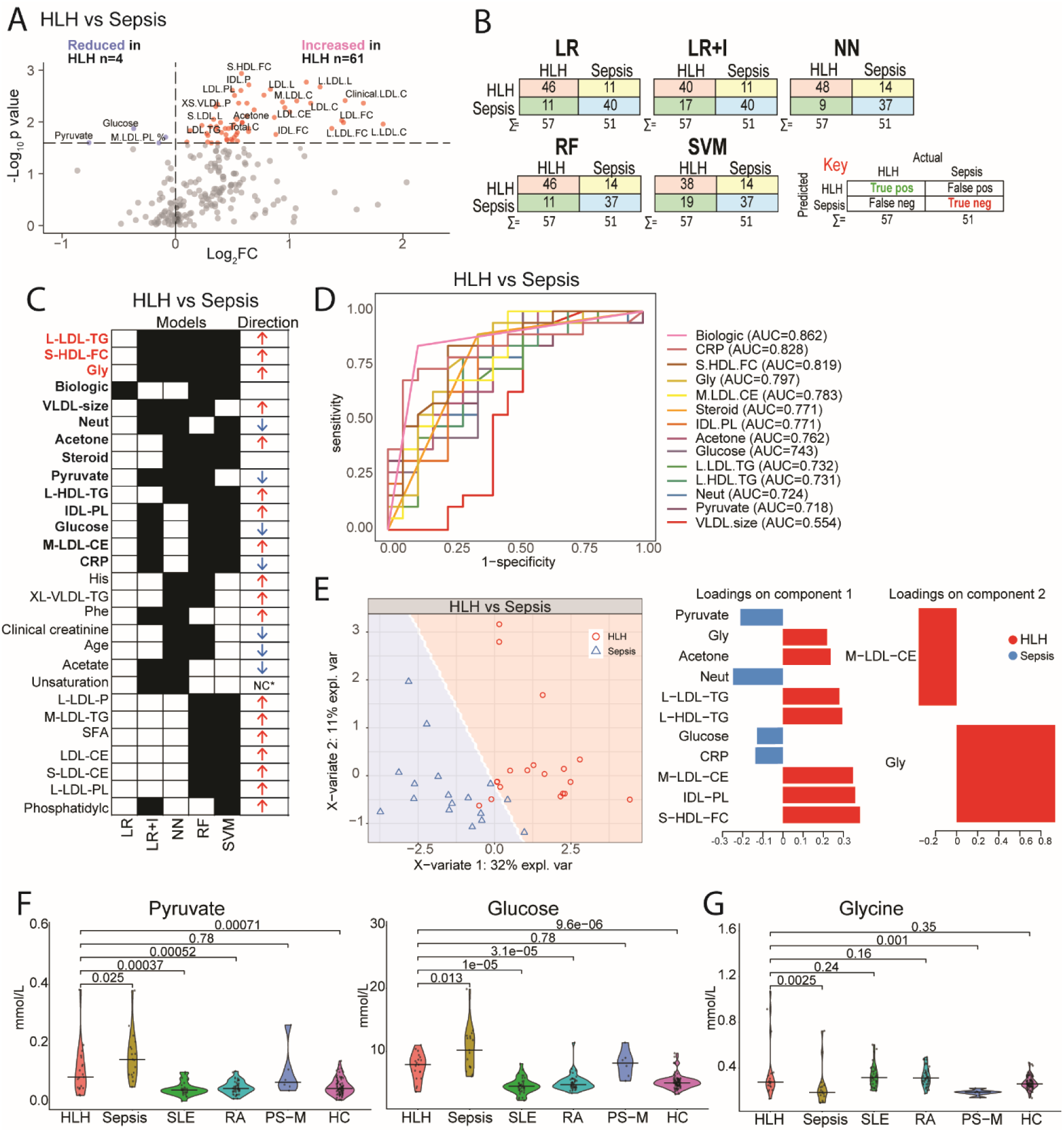
Adults hospitalised with HLH can be distinguished from sepsis using metabolomic and laboratory markers. **(A**) Sera from adults hospitalised with HLH (n=19) and sepsis (n=17), were analysed using an NMR metabolomic platform. Volcano plot shows differentially abundant metabolites in HLH vs. sepsis. Multiple Mann-Whitney U tests, padj<0.1 **(B-E)** Metabolites, demographic and clinical features (bilirubin, creatinine, CRP, lymphocyte, neutrophil, platelet and white cell count and treatment) were analysed by multiple machine learning (ML) models (HLH vs Sepsis): logistic regression (LR) with/without interactions (I), neural network (NN), random forest (RF), support vector machine (SVM). **(B)** Confusion matrices showing the number of correct (red and blue squares) and incorrect (yellow and green squares) classifications for each model. **(C)** Comparison of metabolites selected by each ML model (black squares). Metabolites selected by ≥4 models (red bold); metabolites selected by 2-3 models (black bold), NC= no change. Arrows represent whether the metabolite or clinical feature is elevated (↑) or lower (↓) in HLH vs sepsis. **(D)** Area under the curve-Receiver operator characteristic (AUC-ROC) of top 12 metabolites and/or clinical features in the ML analyses identified in (C) and Figure S3A-B. **(E)** sPLS-DA plot (sparse partial least squares-discriminant analysis) and associated features in components 1 and 2.to validate the metabolomic signature, i.e. top 12 features (excluding treatment) identified in (C) and Figure S3A-B and Table S2 for metabolite abbreviations. **(F-G)** Violin plots showing the abundance of **(F)** glucose, pyruvate and **(G)** glycine in adults with HLH (n=19), sepsis (n=17), SLE (N=37), RA (n=46), post-surgery malignancy (n=6), and healthy controls (n=89). Pairwise comparisons using Mann-Whitney U tests.

Metabolomic data from adults with active rheumatic diseases (SLE, n=37; RA, n=46) attending outpatient clinics (Table 1), were also compared against HLH since they are potential HLH-drivers/triggers. The patterns of metabolic dysregulation were different in patients with chronic inflammation (which can cause HLH) compared to the metabolic dysfunction seen in patients who were acutely unwell with HLH (Table 2 and Table S4, Figure S4A-D). Patients who had undergone major elective surgery for cancer resection (n=6) also demonstrated significant differences in metabolite expression compared to HLH (88 and 39 metabolites were up- and downregulated respectively, padj<0.05) (Figure S4E).

Notably, the HLH-associated metabolites compared to all disease controls (sepsis, SLE and RA) were enriched in metabolic pathways related to lipid metabolism including: “Familial hyperlipidaemia type 1-5”, “SLC-mediated transmembrane transport”, “Lipid particles composition”, “Plasma lipoprotein clearance”, and “Plasma lipoprotein remodelling”, as seen in the HLH vs HC comparison (Table S5). Overall, these results support markedly altered lipoprotein composition in adults with HLH, with a shift toward enrichment of triglycerides and ApoB-containing lipoproteins and depletion of HDL and cholesterol when compared to all control groups.

The results also revealed that glycolysis-related metabolites were reduced in HLH (glucose, pyruvate) compared with sepsis (Figure 4F) but were elevated in HLH compared with chronic inflammatory conditions (SLE, RA) and HCs suggesting distinct differences in energy metabolism and utilisation between HLH and controls. This could contribute to differences in inflammatory and metabolic responses between these conditions. Glycine, an amino acid with a crucial role in metabolic regulation^18^, was elevated in HLH vs sepsis and the surgical cohort supporting a possible compensatory response to increased oxidative stress and immune activation (Figure 4G).

### Metabolomic biomarkers can stratify HLH patients from all disease controls

The potential of metabolites to serve as diagnostic biomarkers for HLH was explored in more detail. Forty-four metabolites stratified adults with HLH from all disease controls (of which 27 achieved an AUC >0.70 across all comparisons) (Figure S5A-C, Table S6). Most of these metabolites were ApoB-containing lipoproteins, triglyceride rich lipoproteins, and saturated fatty acids (SFA). Furthermore, all had statistically significant differences between HLH and control groups and could effectively distinguish HLH from controls either alone or combined as a metabolomic signature (Figure 5A-C, Figure S5D-E for top representative metabolites). These results indicate that metabolites could serve as biomarkers for HLH even in the context of diverse inflammatory and malignant conditions. The shift toward ApoB-containing lipoprotein enrichment in HLH was reinforced when the overall abundance of LDL-particles (P), IDL-P and VLDL-P (non-HDL/ApoB-containing lipoproteins) was examined. All were significantly elevated in HLH vs control groups, whereas HDL-P concentration was significantly dampened in HLH vs controls (except for sepsis which had a trend towards even lower HDL-P levels vs HLH) (Figure 5D).

**Figure 5.**
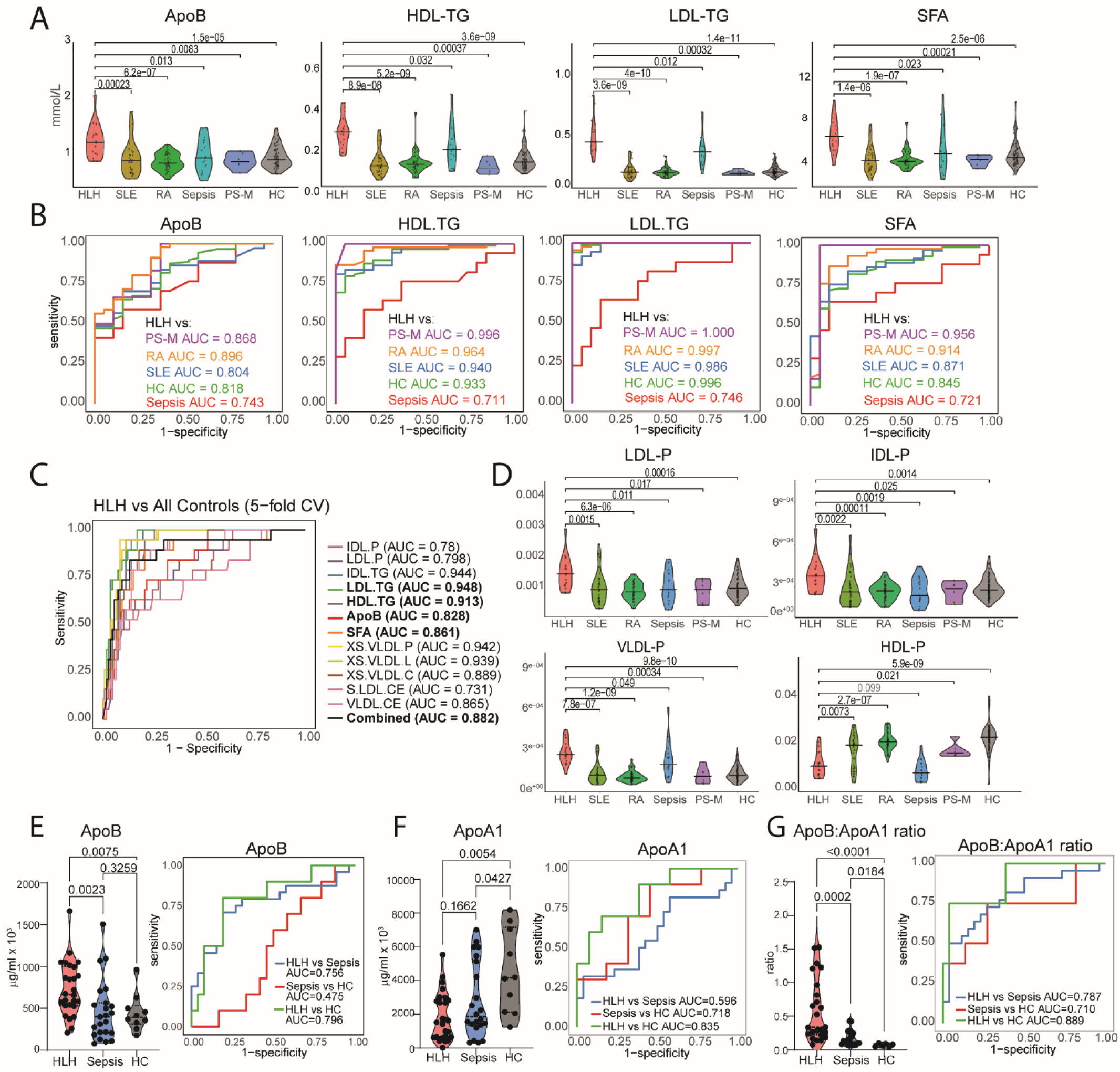
Adults hospitalised with HLH have a distinct metabolite profile compared to patients with sepsis and inflammation controls. **(A)** Violin plots showing pairwise comparisons for selected biomarkers able to stratify HLH from all disease control groups (see Table S6, Figure S5D). P-values computed using Mann-Whitney U tests. **(B)** Multi-receiver operator characteristic (ROC) curves demonstrating how each metabolomic biomarker stratifies HLH from each disease control, area under the curve (AUC) values shown (see Figure S5E). **(C)** Multi-ROC curves showing the performance of representative biomarkers (individually and combined) at stratifying HLH patients from pooled disease controls groups. (**D**) Violin plots showing the abundance of low density, intermediate density, very-low density, and high-density lipoprotein (LDL, IDL, VLDL, HDL) particles (P) Pairwise comparisons computed using Mann-Whitney U tests. (**E-G)** Serum from adults hospitalised with HLH (n=26), sepsis (n=22), and HCs (n=10) were analysed by ELISA for **(E)** Apolipoprotein (Apo)B and **(F)** ApoA1. Data shown as Violin plots with p values from Kruskal-Wallis test and multi-ROC curves and AUC values, **(G)** Violin plot showing ApoB:ApoA1 ratio calculated from data in (E-F) and multi-ROC curves showing AUC values.

Since apolipoprotein measurements are readily available as clinical laboratory assays, the discriminative ability of ApoB, ApoA1 and ApoB:ApoA1 between adults with HLH and sepsis were validated using ELISA methodology. ApoB was significantly elevated in HLH compared to both sepsis (p=0.0023, AUC=0.756) and HCs (p=0.0075, AUC=0.796) (Figure 5E), whereas ApoA1 was reduced in both HLH (p=0.0054, AUC=0.835) and sepsis (p=0.0427, AUC=0.718) compared to HCs (Figure 5F), recapitulating the metabolomic data. Interestingly, the ApoB/ApoA1 ratio was significantly elevated in HLH compared with sepsis (p=0.0002, AUC=0.787) and healthy controls (p<0.0001, AUC=0.710) (Figure 5G). This association remained significant after adjustment for corticosteroid treatment using logistic regression, with each doubling of ApoB/ApoA1 ratio associated with a twofold increase in the likelihood of HLH compared with sepsis (OR=1.9977, 95% CI=1.1562-3.4516, p=0.0131).

To identify potential cut-points for differential diagnosis, ROC analysis was performed to discriminate HLH (n=26) from sepsis (n=22) (Table S7). The ApoB/ApoA1 ratio showed the strongest discrimination (AUC=0.7867), while ApoB alone performed comparably (AUC=0.7465), suggesting that although ApoB/ApoA1 ratio and ApoB are unlikely to function as standalone diagnostic tests, the apolipoprotein dysregulation they reflect may add value when combined with existing clinical tools such as the HScore to improve discrimination between HLH and sepsis.

## DISCUSSION

This study is the first detailed characterisation of the serum metabolome in adults hospitalised with HLH. We report dysregulated serum lipid metabolism that extended beyond reported cases of dampened HDL-C, LDL-C, total cholesterol and hypertriglyceridemia^7–10^. We found that dyslipidaemia in HLH was associated with elevation of ApoB-containing lipoproteins; depletion of HDL particles and ApoA1; and lipoprotein remodelling towards triglyceride enrichment and cholesterol depletion. The metabolomic signature demonstrated a more severe form of dyslipidaemia when compared to sepsis^12^ and chronic inflammatory conditions such as RA^4,19^ and SLE^4,20–22^, where dyslipidaemia is well characterised. Additional metabolic disturbances included changes in glucose homeostasis and amino acid metabolism which could shed light on the pathogenesis of HLH. Finally, by comparing differentially expressed metabolites between adults with HLH and relevant disease control groups, we identified potential HLH-associated biomarkers, which if validated could help to distinguish HLH from HLH-mimics, an important unmet clinical need.

One of the consistent findings in HLH compared to both healthy and disease control groups was abnormal lipoprotein remodelling towards triglyceride enrichment and cholesterol depletion in HDL, VLDL, LDL and IDL. Depletion of HDL particles reduces the lipidation of ApoA1 via lipid transporters^23^. This compromises reverse cholesterol transport (RCT), the process by which excess cholesterol is exported to HDL and delivered to the liver for excretion. Impaired RCT can promote cholesterol accumulation within immune cells, including macrophages, contributing to foam cell formation and exacerbating inflammation^4^. It can also impact immune cell function by influencing immune synapse formation, cell signalling and function including altered cytolytic behaviour^24–27^. Lipoprotein remodelling is also regulated via cholesterol ester transfer protein (CEPT) which transfers cholesteryl esters from HDL to ApoB-containing particles and triglycerides to HDL in a bidirectional manner^28,29^. While CETP gene expression was not significantly altered in HLH compared to HCs, genes associated with N-glycan biosynthesis and quality control were upregulated in HLH. Increased N-glycosylation could contribute to hyperactivated CETP and enhanced subsequent lipoprotein remodelling. In contrast, PLTP (Phospholipid Transfer Protein) was significantly upregulated in HLH vs HCs. PLTP facilitates the bidirectional transfer of phospholipids, free cholesterol, and sphingomyelin between lipoproteins and plays a central role in HDL remodelling^30^. Although PLTP expression increases during inflammation and hypertriglyceridaemia^31^, PLTP hyperactivity destabilises HDL particles, reduces HDL-C levels and accelerates HDL catabolism^30^. Thus, hyperactivation of either CETP or PLTP could accelerate HDL clearance and reduce cholesterol-rich HDL in the serum, contributing to the dysfunctional lipoprotein phenotype we have described in HLH. Lipoprotein remodelling was also altered between adults with HLH and disease control groups, including sepsis. These results are supported by a recent proteomic study^32^, showing ‘plasma lipoprotein remodelling’ was an enriched functional cluster among the 194 proteins differentially expressed between HLH and non-HLH patients^32^. Lipoprotein remodelling is likely to represent a distinguishing metabolic feature between HLH and other hyperinflammatory conditions.

Abnormal lipoprotein profiles could also, at least in part, be driven by inflammatory cytokines including TNF-α and IL-6, both known to be elevated in HLH^1^. However, the lipoprotein signature of HLH was different to patients with RA and SLE, that also have elevated TNF-α and IL-6 during active disease, suggesting other drivers of dyslipidaemia in HLH.

Beyond lipid disturbances, adults with HLH exhibited marked alterations in energy and amino acid metabolism compared to healthy controls. The combination of elevated glucose, lactate, pyruvate, and glycerol, and reduced citrate, suggests a shift toward elevated levels of glycolysis and gluconeogenic substrate mobilisation (e.g. glycerol, lactate, glycine, histidine, valine). However, reduced citrate levels could be associated with increased fatty acid synthesis supported by elevated fatty acid levels in HLH serum, coupled with elevated expression of *IDH1* (isocitrate dehydrogenase-1) which has a dual role in the TCA cycle and fatty acid β-oxidation^33^. A potential source for elevated serum amino acids in HLH could be through processes such as haemophagocytosis or cytokine-mediated eryptosis, leading to the release and breakdown of haemoglobin to haem and globin^34,35^. The globin component is further catabolised into various amino acids^36^ found to be elevated in HLH. As a result, these amino acids could be used for energy synthesis and metabolic pathways including glycolysis and gluconeogenesis.

Compared to sepsis, we show that HLH had lower serum concentrations of glucose and pyruvate, whilst lactate levels remained comparable between the two groups despite the use of steroids. This metabolic profile suggests differences in immunometabolism between HLH and sepsis. In HLH, the persistent activation of cytotoxic lymphocytes and macrophages likely drives sustained glucose uptake and glycolytic flux, resulting in depletion of glucose and pyruvate compared to sepsis, while producing lactate. The preservation of serum lactate despite low levels of upstream metabolites points towards a reliance on glycolysis. Supporting this, Chen et al. (2024) reported increased glucose uptake in T cells and monocytes and increased lipid accumulation in monocytes, NK cells, and T cells from secondary HLH patients^37^. Our gene-metabolite interaction network showed significant upregulation of glucose transporter genes, including *GLUT1*, *GLUT4*, and *GLUT5* and monocarboxylate transporter 1 (*MCT1*), which transport lactate and pyruvate across the plasma membrane^38^ indicating enhanced cellular glucose uptake. Sepsis is often characterised by stress-induced hyperglycaemia^39^, caused by the release of stress hormones including cortisol and insulin counter-regulatory hormones (catecholamines/glucagon/growth hormone) which stimulate gluconeogenesis and glycogenolysis^40,41^. Lactate levels in both conditions were comparable; this could suggest changes in lactate production (glycolysis) and/or consumption (oxphos) between the two conditions. Glycine levels were also significantly higher in adult HLH compared to sepsis consistent with previous studies reporting reduced glycine concentrations in both plasma and liver tissue during sepsis, alongside downregulation of genes involved in glycine biosynthesis and decreased glutathione levels^42,43^. Elevated glycine in HLH may reflect a compensatory response to increased oxidative stress and immune activation and a shift in metabolic programming. Further investigation into metabolic pathways intersecting with glycine metabolism^43^, could reveal further discriminatory features between HLH and sepsis.

While this study is the first to perform a comprehensive serum metabolomic analysis of HLH we acknowledge some limitations. Metabolomic measurements can be influenced by factors such as age, diet^44^, and hormonal status^45^ and whilst NMR-based metabolomics (used within this study) is more cost effective, reproducible, and requires minimal sample preparation compared to mass spectroscopy methods, the number of molecules measured by NMR is more limited^46^. Sample sizes were unbalanced between HLH patients compared to healthy and disease controls. This was mitigated using ML analysis through down-sampling and repeated five-fold cross-validation. Differences in demographic information were seen between HLH and disease controls particularly with age and sex and all patients had received either steroid or disease modifying therapy which are known to influence serum lipid levels^47^, this was accounted for in the ML models and univariate logistic regression analyses, where we demonstrated that metabolites alone can stratify HLH patients from these various controls.

In conclusion, this study provides evidence of systemic metabolic disturbances that span the transcriptome and metabolome in adults with HLH. Notably, adults with HLH demonstrated dyslipidaemia and distinct patterns of lipoprotein remodelling, which may reflect underlying disease mechanisms and could serve as potential treatment targets or biomarkers to discriminate HLH from other inflammatory conditions.

## Data Availability

All data produced in the present study are available upon reasonable request to the authors

## Data sharing statement

For original data please contact.

## ACKNOWLEDGEMENTS

Sample collection for sepsis and surgical patients was coordinated by Dr David Breaely (UCLH).

## AUTHORSHIP CONTRIBUTIONS

Designed and supervised the research; ECJ, AEO, JJM, GAR, TM

Recruited patients: JJM, AH, NA, KL, LR, AEO.

Curated patient databases: AEO, AH, LR, JJM, NA

Performed metabolomic and transcriptomic analysis, designed and performed machine learning analysis: AEO

Performed ELISAs: KL, AEO

Wrote the manuscript: AEO, ECJ

Critical review: JJM, GAR, TM, LR, KL, NA

