## Supplementary Information for "Lipoprotein remodelling and elevated serum apolipoprotein B distinguish adults with haemophagocytic lymphohistiocytosis from sepsis"

**Supplementary Methods:**

**Patient and control cohorts** (see Table 1)

*SLE:* Serum from patients with SLE (n=37) diagnosed according to the American College of Rheumatology classification for SLE or the Systemic Lupus International Collaborating Clinics criteria recruited from UCLH Rheumatology outpatient clinics, had active disease assessed using Systemic Lupus Erythematosus Disease Activity Index (SLEDAI) score ≥6 or British Isles Lupus Assessment Group (BILAG) score ≥8[16].

*RA:* Serum from patients with RA (n=46) diagnosed according to the European League Against Rheumatism/American College of Rheumatology 2010 criteria and referred to the UCLH rheumatologist-led ultrasound outpatient clinic with ultrasound-confirmed synovitis in at least one joint positive for Power Doppler signal, disease activity was assessed using disease activity score-28-C reactive protein (DAS28-CRP, active disease >3.2).

*Healthy controls:* Serum from healthy volunteers (HC, n=89) obtained from volunteers within UCL.

*Ethical Approval***:** All participants were recruited after gaining informed written consent. Healthy volunteers were recruited under London-Harrow Research Ethics approval (11/LO/0330). Patients with SLE were recruited under London-Harrow Research Ethics approval (11/LO/0330). Patients with RA were recruited with approval from the Hampstead Research Ethics Committee (14/LO/1506).

**Metabolomics**

This metabolomic platform provides clinical-grade analysis that is fast, cost-effective, and reproducible. Sera not exposed to freeze/thaw cycles were analysed. The success rate of biomarker quantification was >99% across the cohorts, and metabolite concentrations fell within the distributions commonly observed in general population cohorts(1).

**Data handling and analysis**

*Imputation:* Missing values were imputed using kNN using a default value of *k*=5 within the feature space of R package bnstruct.

*Homology reduction:* Since many metabolites were biologically interdependent, and highly correlated only ratios and absolute concentrations were used for ML models to reduce dimensionality and multicollinearity (n=172 metabolites). Homology reduction was applied whereby if two features had a correlation coefficient >0.95, then the feature with the greatest mean absolute correlation with the remaining features was removed(1). The final list of features was used within the machine learning (ML) analysis.

*Predictors:* Homology-reduced datasets included in ML were as follows: HLH vs. HC (73 metabolites), HLH vs. sepsis (92 metabolites), HLH vs. SLE (63 metabolites), HLH vs. RA (78 metabolites). In addition cohort information was included where available: age, sex, ethnicity, treatment, sodium (Na), potassium (K), creatinine, eGFR, ALT, albumin, alkaline phosphatase (ALP), bilirubin, triglyceride, ferritin, CRP, haemoglobin, white cell count (WCC), neutrophil, lymphocyte, platelets, red cell count (RCC), erythrocyte sedimentation rate (ESR) and international normalised ratio (INR).

*Data scaling*: Metabolites were centred on the mean and scaled to the standard deviation(1).

on the best-performing ML model.

**Predictive models:**

*Logistic regression with/without interactions:* Less important features were shrunk to zero, using the least absolute shrinkage and selection operator (lasso) method, which uses the absolute value of the coefficient as a penalty. Tuning the regularisation variable lambda (λ), determines the strength of shrinkage(1). Lasso logistic regression (including with and without interactions) was conducted using the glmnet R package(3).

*eXtreme Gradient Boosting:* XGBoost is a supervised machine learning algorithm that builds an ensemble of decision trees using a technique called gradient boosting. Rather than training a single complex model, XGBoost sequentially constructs a series of simpler decision trees, with each new tree focusing on correcting the errors made by the previous ones. Over time, the model improves by placing more emphasis on samples, i.e., patients that were previously misclassified. This approach allows the model to better handle difficult-to-predict cases. It does this whilst incorporating regularisation techniques to reduce the risk of overfitting. By combining the outputs of multiple weak learners/decision trees, XGBoost can achieve stronger predictive performance than singular ML models(4). Model training involved ten-fold cross-validation and parameter tuning with the following settings: sampling = "down", nrounds = 100, 200, max_depth = 10, 15, 20, 25, colsample_bytree = seq(0.5, 0.9, length.out = 5), eta = 0.1, gamma = 0, min_child_weight = 1, and subsample = 1. Feature importance was evaluated using information gain, which measures the contribution of each variable to improving model performance.

*Random forest:* RF is a machine-learning algorithm that assigns observations into classes (hosp-HLH/recov-HLH/HCs/disease controls) by creating thousands of decision trees (predicts the value of target variables by learning simple decision rules inferred from the features, i.e., metabolites from the dataset), or a "forest", and averaging the results. Only a small random sample of predictors are candidates for selection at each node, so the created trees are decorrelated. Importance was quantified by the Gini index, which represents the total variance across the two classes, the quality of each split and the purity of each node(1). The values for mtry and ntree were tuned using the R Package caret(5). The parameters were set to mtry = 10, ntree = 1,000.

*Support vector machine:* SVM is a supervised classification method which optimally separates data into two classes by creating a hyperplane. The radial basis function kernel was used, if the dataset for a given comparison was not linearly separable. Values for C, epsilon and gamma were tuned using the R Package caret(5). The parameters were set to C = 4.0, epsilon = 0.1, gamma = 0.01(1).

*Neural network:* NN uses the multi-layer perceptron algorithm with backpropagation, which can learn both linear and non-linear models. NNs incorporate large numbers of processing nodes that are densely interconnected, similar to the human brain. These nodes are organised into layers-input, hidden and output.

The output node/classification is affected by weights of values in hidden layers; these weights are adjusted when the model is trained for the best classification(1).

**Model performance:**

Model performance metrics were calculated using RStudio 4.4.2 (The R Foundation, Vienna, Austria) based on predictions across all cross-validation test folds (five-fold cross-validation repeated three times). This approach was chosen due to modest sample sizes in each binary classification comparison. Repeating the cross-validation three times helps reduce variability due to random fold assignments, thus helping to stabilise and make estimates of model performance more reliable. As such, the total number of predictions exceeds the number of unique samples. The following performance metrics were recorded: sensitivity/recall (the true positive rate), specificity (the true negative rate), classification accuracy (the proportion of correctly classified cases), AUC-ROC(1) and Matthew's Correlation Coefficient (MCC, a comprehensive measure of classification quality that accounts for all parts of the confusion matrix, including true and false positives and negatives). MCC was used to determine the best-performing ML model for each comparison, as it provides a balanced evaluation even when class sizes are unequal, or sample sizes are small. Unlike accuracy, MCC is not biased by class imbalance and offers a more informative single metric that reflects overall model quality. Only models with an MCC or AUC greater than 0.40 and 0.70, respectively, were considered for downstream analysis.

To visualise how well individual metabolites (identified by six ML models) could discriminate HLH patients from HCs and disease controls, compared to the metabolomic signature, AUC-ROC curves were generated based on the best-performing ML model. RStudio 4.4.2 (The R Foundation, Vienna, Austria) was used for ML analysis.

**Sparse Partial Least Squares Discriminant Analysis (sPLS-DA):** The identified metabolomic signature for each comparison was validated through sPLS-DA, a supervised clustering ML approach that combines classification and parameter selection into one operation using the R package *mixOmics*(6). sPLS-DA models with different component numbers were assessed by 10-fold cross-validation and 50 repetitions using the balanced error rate to evaluate model performance. The number of components were selected based on the combination that gave the lowest overall estimation error rate. These were selected as optimal, giving the best discriminatory performance for further analysis. The separation of the comparisons was presented by projecting the samples into the subspace constructed by component 1 and component 2(1). The top weighted features were selected and presented through variable loading plots. sPLS-DA results were visualised using the R package ggplot2(7).

**Metabolite set enrichment analysis:** Performed on the metabolomic signature identified by ML models (all metabolites identified by at least one ML model) and multiple Mann-Whitney U tests, using the RaMP-DB database (Relational Database of Metabolomics Pathways) within MetaboAnalyst *6.0*[18]. This generated a report on over-representation analysis. Only metabolic pathways FDR<0.05 were considered.

**Transcriptomic analysis**

***RNA preparation and sequencing**:* Whole blood RNA was isolated by UCL Genomics from adults hospitalised, patients with HLH (n=19) and HCs (n=14) using the Qiagen PAXgene Blood RNA extraction kit. Sample concentration and purity were determined using a NanoDropTM 1000 Spectrophotometer. UCL Genomics performed sequencing, quality control analysis, genome alignment, and quantification from 50 ng of isolated RNA/sample (London, UK). Illumina technology was used to produce high-throughput sequencing data. This data was processed by Picard(8), aligned to the GRCH38 homo sapiens reference genome by STAR(9), and processed and filtered by fastp (1, 10).

***Differential gene expression analysis**:* Differential gene expression was determined by DESeq2 (Version: 1.46.0) analysis using Wald hypothesis testing and parametric fit typing, and using the Benjamin-Hochberg method for multiple test correction(11). No genes were removed using the DESeq2 filtering process, which made the p-value adjustment more stringent. For the hosp-HLH vs recov-HLH comparison, whilst the two patient cohorts were age and ethnicity matched, there were statistically significant differences in sex. To account for this, the design formula was modified to control for sex while testing for condition and the adjusted and unadjusted results were compared against each other to ensure the core pathways were maintained and not driven by sex differences. Using the entirety of the gene set, principal component analysis (PCA) was used to reduce the dimensionality of the dataset and to visualise the level of clustering and separation between: hosp-HLH vs. recov-HLH; hosp-HLH vs. HCs; recov-HLH vs. HCs. Lastly, using a reduced number of genes, after applying a threshold of ≥25 counts, volcano plots were generated to visualise the differential expression analysis results. As well as to identify the number of genes which passed a threshold of both FDR<0.05 and FC±1.5 (log2FC: 0.585)(1).

***Metabolic module analysis**:* To validate the metabolomic findings, metabolic module analysis was conducted using Metabolizer (Metabolizer) on differentially expressed gene (DEG) datasets comparing adults hospitalised with HLH (n=19) to HCs (n=14) - *all samples had matching metabolomic analysis performed on serum from the same day*. This approach identified metabolic pathways associated with DEGs and had the benefit of additionally identifying pathways not represented/covered by the metabolomic panel. Pathways with significant enrichment (p<0.05) were recorded, along with the direction of regulation based on up- and downregulated genes. DEGs associated with each metabolic module/pathway were manually extracted using the KEGG module database (KEGG MODULE Database).

**ELISA**

Serum levels of ApoB and ApoA1 in adults with HLH were validated using ELISA (Quantakine, Biotechne). Assays were performed in a larger cohort of adults hospitalised with HLH (n=26), sepsis (n=22), and HCs (n=10) as per manufacturer's instructions, except for using a 1/2000 dilution in the ApoB ELISA and 1/25000 in the ApoA1 ELISA. Fluorescence was measured using TECAN Spark Multimode Plate Reader at 450nm absorbance with a correction wavelength of 540nm.

*Optimum apolipoprotein cut-offs:* To identify the optimal ApoA1, ApoB and ApoB/ApoA1 threshold for distinguishing between HLH and Sepsis patient groups, the Youden Index method was used(254). The Youden Index maximises the sum of sensitivity and specificity to identify the point on the receiver operating characteristic (ROC) curve that best separates the two classes. ELISA measurements were used as the predictor variable, and disease group classification (HLH/Sepsis) was used as the binary outcome. To assess the robustness and stability of the cut-off, non-parametric bootstrapping with 10,000 resamples was performed. The output included the optimal cutoff point, confidence intervals, the corresponding sensitivity, specificity and AUC. This was done using the R package *cutpointr*.

**Table S1: Clinical characteristics and treatment of adults hospitalised with HLH (n=19)**

| | | |
|---|---|---|
| | *Infection, n (%)* | 5 (26.32) |
| | *Malignancy (%)* | 8 (42.11) |
| **Primary trigger** | *Rheumatological (%)* | 3 (15.79) |
| | *Genetic (%)* | 1 (5.26) |
| | *Unknown/Other (%)* | 2 (10.53) |
| | *Malignancy, n (%)* | 3 (15.79) |
| | *Rheumatological, n (%)* | 0 (0.00) |
| **Risk factors** | *Other infection (%)* | 1 (5.26) |
| | *Genetic (%)* | 1 (5.26) |
| | *Unknown/Other (%)* | 3 (15.79) |

**Table S2: Full list of serum metabolites measured by the Nightingale Health NMR spectroscopy platform.** This metabolomic platform provides clinical-grade analysis that is fast, cost-effective, and reproducible. Sera not exposed to freeze/thaw cycles were analysed. The success rate of biomarker quantification was >99% across the cohorts, and metabolite concentrations fell within the distributions commonly observed in general population cohorts(1).

| Abbreviation | Full name |
|---|---|
| **Total-C** | Total cholesterol |
| **non-HDL-C** | Total cholesterol minus HDL-C |
| **Remnant-C** | Remnant cholesterol (non-HDL, non-LDL cholesterol) |
| **VLDL-C** | VLDL cholesterol |
| **Clinical LDL-C** | Clinical LDL cholesterol |
| **LDL-C** | LDL cholesterol |
| **HDL-C** | HDL cholesterol |
| **Total-TG** | Total triglycerides |
| **VLDL-TG** | Triglycerides in VLDL |
| **LDL-TG** | Triglycerides in LDL |
| **HDL-TG** | Triglycerides in HDL |
| **Total-PL** | Total phospholipids in lipoprotein particles |
| **VLDL-PL** | Phospholipids in VLDL |
| **LDL-PL** | Phospholipids in LDL |
| **HDL-PL** | Phospholipids in HDL |
| **Total-CE** | Total esterified cholesterol |
| **VLDL-CE** | Cholesteryl esters in VLDL |
| **LDL-CE** | Cholesteryl esters in LDL |
| **HDL-CE** | Cholesteryl esters in HDL |
| **Total-FC** | Total free cholesterol |
| **VLDL-FC** | Free cholesterol in VLDL |
| **LDL-FC** | Free cholesterol in LDL |
| **HDL-FC** | Free cholesterol in HDL |
| **Total-L** | Total lipids in lipoprotein particles |
| **VLDL-L** | Total lipids in VLDL |
| **LDL-L** | Total lipids in LDL |
| **HDL-L** | Total lipids in HDL |
| **Total-P** | Total concentration of lipoprotein particles |

| Abbreviation | Full name |
|---|---|
| **VLDL-P** | Concentration of VLDL particles |
| **LDL-P** | Concentration of LDL particles |
| **HDL-P** | Concentration of HDL particles |
| **VLDL size** | Average diameter for VLDL particles |
| **LDL size** | Average diameter for LDL particles |
| **HDL size** | Average diameter for HDL particles |
| **Phosphoglyc** | Phosphoglycerides |
| **TG/PG** | Ratio of triglycerides to phosphoglycerides |
| **Cholines** | Total cholines |
| **Phosphatidylc** | Phosphatidylcholines |
| **Sphingomyelins** | Sphingomyelins |
| **ApoB** | Apolipoprotein B |
| **ApoA1** | Apolipoprotein A1 |
| **ApoB/ApoA1** | Ratio of apolipoprotein B to apolipoprotein A1 |
| **Total-FA** | Total fatty acids |
| **Unsaturation** | Degree of unsaturation |
| **Omega-3** | Omega-3 fatty acids |
| **Omega-6** | Omega-6 fatty acids |
| **PUFA** | Polyunsaturated fatty acids |
| **MUFA** | Monounsaturated fatty acids |
| **SFA** | Saturated fatty acids |
| **LA** | Linoleic acid |
| **DHA** | Docosahexaenoic acid |
| **Omega-3 %** | Ratio of omega-3 fatty acids to total fatty acids |
| **Omega-6 %** | Ratio of omega-6 fatty acids to total fatty acids |

| Abbreviation | Full name |
|---|---|
| **PUFA %** | Ratio of polyunsaturated fatty acids to total fatty acids |
| **MUFA %** | Ratio of monounsaturated fatty acids to total fatty acids |
| **SFA %** | Ratio of saturated fatty acids to total fatty acids |
| **LA %** | Ratio of linoleic acid to total fatty acids |
| **DHA %** | Ratio of docosahexaenoic acid to total fatty acids |
| **PUFA/MUFA** | Ratio of polyunsaturated fatty acids to monounsaturated fatty acids |
| **Omega-6/Omega-3** | Ratio of omega-6 fatty acids to omega-3 fatty acids |
| **Ala** | Alanine |
| **Gln** | Glutamine |
| **Gly** | Glycine |
| **His** | Histidine |
| **Total BCAA** | Total concentration of branched-chain amino acids (leucine + isoleucine + valine) |
| **Ile** | Isoleucine |
| **Leu** | Leucine |
| **Val** | Valine |
| **Phe** | Phenylalanine |
| **Tyr** | Tyrosine |
| **Glucose** | Glucose |
| **Lactate** | Lactate |
| **Pyruvate** | Pyruvate |
| **Citrate** | Citrate |
| **Glycerol** | Glycerol |
| **bOHbutyrate** | 3-Hydroxybutyrate |
| **Acetate** | Acetate |
| **Acetoacetate** | Acetoacetate |
| **Acetone** | Acetone |
| **Creatinine** | Creatinine |

| Abbreviation | Full name |
|---|---|
| **Albumin** | Albumin |
| **GlycA** | Glycoprotein acetyls |
| **XXL-VLDL-P** | Concentration of chylomicrons and extremely large VLDL particles |
| **XXL-VLDL-L** | Total lipids in chylomicrons and extremely large VLDL |
| **XXL-VLDL-PL** | Phospholipids in chylomicrons and extremely large VLDL |
| **XXL-VLDL-C** | Cholesterol in chylomicrons and extremely large VLDL |
| **XXL-VLDL-CE** | Cholesteryl esters in chylomicrons and extremely large VLDL |
| **XXL-VLDL-FC** | Free cholesterol in chylomicrons and extremely large VLDL |
| **XXL-VLDL-TG** | Triglycerides in chylomicrons and extremely large VLDL |
| **XL-VLDL-P** | Concentration of very large VLDL particles |
| **XL-VLDL-L** | Total lipids in very large VLDL |
| **XL-VLDL-PL** | Phospholipids in very large VLDL |
| **XL-VLDL-C** | Cholesterol in very large VLDL |
| **XL-VLDL-CE** | Cholesteryl esters in very large VLDL |
| **XL-VLDL-FC** | Free cholesterol in very large VLDL |
| **XL-VLDL-TG** | Triglycerides in very large VLDL |
| **L-VLDL-P** | Concentration of large VLDL particles |
| **L-VLDL-L** | Total lipids in large VLDL |

| Abbreviation | Full name |
|---|---|
| **L-VLDL-PL** | Phospholipids in large VLDL |
| **L-VLDL-C** | Cholesterol in large VLDL |
| **L-VLDL-CE** | Cholesteryl esters in large VLDL |
| **L-VLDL-FC** | Free cholesterol in large VLDL |
| **L-VLDL-TG** | Triglycerides in large VLDL |
| **M-VLDL-P** | Concentration of medium VLDL particles |
| **M-VLDL-L** | Total lipids in medium VLDL |
| **M-VLDL-PL** | Phospholipids in medium VLDL |
| **M-VLDL-C** | Cholesterol in medium VLDL |
| **M-VLDL-CE** | Cholesteryl esters in medium VLDL |
| **M-VLDL-FC** | Free cholesterol in medium VLDL |
| **M-VLDL-TG** | Triglycerides in medium VLDL |
| **S-VLDL-P** | Concentration of small VLDL particles |
| **S-VLDL-L** | Total lipids in small VLDL |
| **S-VLDL-PL** | Phospholipids in small VLDL |
| **S-VLDL-C** | Cholesterol in small VLDL |
| **S-VLDL-CE** | Cholesteryl esters in small VLDL |
| **S-VLDL-FC** | Free cholesterol in small VLDL |
| **S-VLDL-TG** | Triglycerides in small VLDL |
| **XS-VLDL-P** | Concentration of very small VLDL particles |
| **XS-VLDL-L** | Total lipids in very small VLDL |

| Abbreviation | Full name |
|---|---|
| **XS-VLDL-PL** | Phospholipids in very small VLDL |
| **XS-VLDL-C** | Cholesterol in very small VLDL |
| **XS-VLDL-CE** | Cholesteryl esters in very small VLDL |
| **XS-VLDL-FC** | Free cholesterol in very small VLDL |
| **XS-VLDL-TG** | Triglycerides in very small VLDL |
| **IDL-P** | Concentration of IDL particles |
| **IDL-L** | Total lipids in IDL |
| **IDL-PL** | Phospholipids in IDL |
| **IDL-C** | Cholesterol in IDL |
| **IDL-CE** | Cholesteryl esters in IDL |
| **IDL-FC** | Free cholesterol in IDL |
| **IDL-TG** | Triglycerides in IDL |
| **L-LDL-P** | Concentration of large LDL particles |
| **L-LDL-L** | Total lipids in large LDL |
| **L-LDL-PL** | Phospholipids in large LDL |
| **L-LDL-C** | Cholesterol in large LDL |
| **L-LDL-CE** | Cholesteryl esters in large LDL |
| **L-LDL-FC** | Free cholesterol in large LDL |
| **L-LDL-TG** | Triglycerides in large LDL |
| **M-LDL-P** | Concentration of medium LDL particles |
| **M-LDL-L** | Total lipids in medium LDL |
| **M-LDL-PL** | Phospholipids in medium LDL |
| **M-LDL-C** | Cholesterol in medium LDL |
| **M-LDL-CE** | Cholesteryl esters in medium LDL |
| **M-LDL-FC** | Free cholesterol in medium LDL |

| Abbreviation | Full name |
|---|---|
| **M-LDL-TG** | Triglycerides in medium LDL |
| **S-LDL-P** | Concentration of small LDL particles |
| **S-LDL-L** | Total lipids in small LDL |
| **S-LDL-PL** | Phospholipids in small LDL |
| **S-LDL-C** | Cholesterol in small LDL |
| **S-LDL-CE** | Cholesteryl esters in small LDL |
| **S-LDL-FC** | Free cholesterol in small LDL |
| **S-LDL-TG** | Triglycerides in small LDL |
| **XL-HDL-P** | Concentration of very large HDL particles |
| **XL-HDL-L** | Total lipids in very large HDL |
| **XL-HDL-PL** | Phospholipids in very large HDL |
| **XL-HDL-C** | Cholesterol in very large HDL |
| **XL-HDL-CE** | Cholesteryl esters in very large HDL |
| **XL-HDL-FC** | Free cholesterol in very large HDL |
| **XL-HDL-TG** | Triglycerides in very large HDL |
| **L-HDL-P** | Concentration of large HDL particles |
| **L-HDL-L** | Total lipids in large HDL |
| **L-HDL-PL** | Phospholipids in large HDL |
| **L-HDL-C** | Cholesterol in large HDL |
| **L-HDL-CE** | Cholesteryl esters in large HDL |
| **L-HDL-FC** | Free cholesterol in large HDL |
| **L-HDL-TG** | Triglycerides in large HDL |
| **M-HDL-P** | Concentration of medium HDL particles |

| Abbreviation | Full name |
|---|---|
| **M-HDL-L** | Total lipids in medium HDL |
| **M-HDL-PL** | Phospholipids in medium HDL |
| **M-HDL-C** | Cholesterol in medium HDL |
| **M-HDL-CE** | Cholesteryl esters in medium HDL |
| **M-HDL-FC** | Free cholesterol in medium HDL |
| **M-HDL-TG** | Triglycerides in medium HDL |
| **S-HDL-P** | Concentration of small HDL particles |
| **S-HDL-L** | Total lipids in small HDL |
| **S-HDL-PL** | Phospholipids in small HDL |
| **S-HDL-C** | Cholesterol in small HDL |
| **S-HDL-CE** | Cholesteryl esters in small HDL |
| **S-HDL-FC** | Free cholesterol in small HDL |
| **S-HDL-TG** | Triglycerides in small HDL |
| **XXL-VLDL-PL %** | Phospholipids to total lipids ratio in chylomicrons and extremely large VLDL |
| **XXL-VLDL-C %** | Cholesterol to total lipids ratio in chylomicrons and extremely large VLDL |
| **XXL-VLDL-CE %** | Cholesteryl esters to total lipids ratio in chylomicrons and extremely large VLDL |
| **XXL-VLDL-FC %** | Free cholesterol to total lipids ratio in chylomicrons and extremely large VLDL |
| **XXL-VLDL-TG %** | Triglycerides to total lipids ratio in chylomicrons and extremely large VLDL |

| Abbreviation | Full name |
|---|---|
| **XL-VLDL-PL %** | Phospholipids to total lipids ratio in very large VLDL |
| **XL-VLDL-C %** | Cholesterol to total lipids ratio in very large VLDL |
| **XL-VLDL-CE %** | Cholesteryl esters to total lipids ratio in very large VLDL |
| **XL-VLDL-FC %** | Free cholesterol to total lipids ratio in very large VLDL |
| **XL-VLDL-TG %** | Triglycerides to total lipids ratio in very large VLDL |
| **L-VLDL-PL %** | Phospholipids to total lipids ratio in large VLDL |
| **L-VLDL-C %** | Cholesterol to total lipids ratio in large VLDL |
| **L-VLDL-CE %** | Cholesteryl esters to total lipids ratio in large VLDL |
| **L-VLDL-FC %** | Free cholesterol to total lipids ratio in large VLDL |
| **L-VLDL-TG %** | Triglycerides to total lipids ratio in large VLDL |
| **M-VLDL-PL %** | Phospholipids to total lipids ratio in medium VLDL |
| **M-VLDL-C %** | Cholesterol to total lipids ratio in medium VLDL |
| **M-VLDL-CE %** | Cholesteryl esters to total lipids ratio in medium VLDL |
| **M-VLDL-FC %** | Free cholesterol to total lipids ratio in medium VLDL |
| **M-VLDL-TG %** | Triglycerides to total lipids ratio in medium VLDL |
| **S-VLDL-PL %** | Phospholipids to total lipids ratio in small VLDL |
| **S-VLDL-C %** | Cholesterol to total lipids ratio in small VLDL |

| Abbreviation | Full name |
|---|---|
| **S-VLDL-CE %** | Cholesteryl esters to total lipids ratio in small VLDL |
| **S-VLDL-FC %** | Free cholesterol to total lipids ratio in small VLDL |
| **S-VLDL-TG %** | Triglycerides to total lipids ratio in small VLDL |
| **XS-VLDL-PL %** | Phospholipids to total lipids ratio in very small VLDL |
| **XS-VLDL-C %** | Cholesterol to total lipids ratio in very small VLDL |
| **XS-VLDL-CE %** | Cholesteryl esters to total lipids ratio in very small VLDL |
| **XS-VLDL-FC %** | Free cholesterol to total lipids ratio in very small VLDL |
| **XS-VLDL-TG %** | Triglycerides to total lipids ratio in very small VLDL |
| **IDL-PL %** | Phospholipids to total lipids ratio in IDL |
| **IDL-C %** | Cholesterol to total lipids ratio in IDL |
| **IDL-CE %** | Cholesteryl esters to total lipids ratio in IDL |
| **IDL-FC %** | Free cholesterol to total lipids ratio in IDL |
| **IDL-TG %** | Triglycerides to total lipids ratio in IDL |
| **L-LDL-PL %** | Phospholipids to total lipids ratio in large LDL |
| **L-LDL-C %** | Cholesterol to total lipids ratio in large LDL |
| **L-LDL-CE %** | Cholesteryl esters to total lipids ratio in large LDL |
| **L-LDL-FC %** | Free cholesterol to total lipids ratio in large LDL |
| **L-LDL-TG %** | Triglycerides to total lipids ratio in large LDL |

| Abbreviation | Full name |
|---|---|
| **M-LDL-PL %** | Phospholipids to total lipids ratio in medium LDL |
| **M-LDL-C %** | Cholesterol to total lipids ratio in medium LDL |
| **M-LDL-CE %** | Cholesteryl esters to total lipids ratio in medium LDL |
| **M-LDL-FC %** | Free cholesterol to total lipids ratio in medium LDL |
| **M-LDL-TG %** | Triglycerides to total lipids ratio in medium LDL |
| **S-LDL-PL %** | Phospholipids to total lipids ratio in small LDL |
| **S-LDL-C %** | Cholesterol to total lipids ratio in small LDL |
| **S-LDL-CE %** | Cholesteryl esters to total lipids ratio in small LDL |
| **S-LDL-FC %** | Free cholesterol to total lipids ratio in small LDL |
| **S-LDL-TG %** | Triglycerides to total lipids ratio in small LDL |
| **XL-HDL-PL %** | Phospholipids to total lipids ratio in very large HDL |
| **XL-HDL-C %** | Cholesterol to total lipids ratio in very large HDL |
| **XL-HDL-CE %** | Cholesteryl esters to total lipids ratio in very large HDL |
| **XL-HDL-FC %** | Free cholesterol to total lipids ratio in very large HDL |
| **XL-HDL-TG %** | Triglycerides to total lipids ratio in very large HDL |
| **L-HDL-PL %** | Phospholipids to total lipids ratio in large HDL |
| **L-HDL-C %** | Cholesterol to total lipids ratio in large HDL |
| **L-HDL-CE %** | Cholesteryl esters to total lipids ratio in large HDL |

| Abbreviation | Full name |
|---|---|
| **L-HDL-FC %** | Free cholesterol to total lipids ratio in large HDL |
| **L-HDL-TG %** | Triglycerides to total lipids ratio in large HDL |
| **M-HDL-PL %** | Phospholipids to total lipids ratio in medium HDL |
| **M-HDL-C %** | Cholesterol to total lipids ratio in medium HDL |
| **M-HDL-CE %** | Cholesteryl esters to total lipids ratio in medium HDL |
| **M-HDL-FC %** | Free cholesterol to total lipids ratio in medium HDL |
| **M-HDL-TG %** | Triglycerides to total lipids ratio in medium HDL |
| **S-HDL-PL %** | Phospholipids to total lipids ratio in small HDL |
| **S-HDL-C %** | Cholesterol to total lipids ratio in small HDL |
| **S-HDL-CE %** | Cholesteryl esters to total lipids ratio in small HDL |
| **S-HDL-FC %** | Free cholesterol to total lipids ratio in small HDL |
| **S-HDL-TG %** | Triglycerides to total lipids ratio in small HDL |

**Supplemental Table S3: List of genes associated with pathways identified from metabolic module analysis for the HLH vs. HC comparison.** Description of gene functions was taken from GeneCards(12). (see Figure 3, Figure S1F)

| Genes | Log$^2$FC | padj | Metabolic modules | Function |
|---|---|---|---|---|
| ENO1 | 0.8861 | 1.25E-05 | Glycolysis, Gluconeogenesis | Catalyses the conversion of 2-phosphoglycerate to phosphoenolpyruvate in glycolysis. |
| GPI | 0.9241 | 1.35E-03 | Glycolysis, Pentose phosphate pathway | Functions as glucose-6-phosphate isomerase, interconverting glucose-6-phosphate and fructose-6-phosphate. |
| FH | 0.5905 | 4.09E-02 | Citrate cycle (TCA cycle, Krebs cycle) | Catalyses the hydration of fumarate to malate in the tricarboxylic acid (TCA) cycle. |
| IDH1 | 0.7827 | 8.01E-04 | Citrate cycle (TCA cycle, Krebs cycle) | Catalyses the oxidative decarboxylation of isocitrate to α-ketoglutarate in the cytoplasm, producing NADPH. |
| KL | 2.0216 | 4.15E-04 | Glucuronate pathway | Encodes the klotho protein, which regulates phosphate and calcium homeostasis and has anti-ageing properties. |
| PHGDH | 1.3914 | 1.69E-03 | Serine biosynthesis | Catalyses the first committed step of serine biosynthesis from 3-phosphoglycerate. |
| ARG1 | 3.0767 | 6.93E-10 | Urea cycle, Polyamine biosynthesis | Converts arginine into urea and ornithine in the urea cycle. |
| BCAT1 | 1.8295 | 4.82E-05 | Leucine degradation | Catalyses reversible transamination of branched-chain amino acids (BCAAs) to their corresponding keto acids. |
| FAH | 1.5413 | 1.56E-04 | Tyrosine degradation | Catalyses the final step in tyrosine catabolism, converting fumarylacetoacetate into fumarate and acetoacetate. |
| NME1 | 1.5244 | 2.75E-04 | Purine metabolism, Pyrimidine ribonucleotide biosynthesis | Functions as a nucleoside diphosphate kinase, maintaining balanced nucleotide pools in the cell. |
| CMPK2 | 1.4001 | 6.15E-03 | Pyrimidine ribonucleotide biosynthesis | Phosphorylates CMP to CDP in mitochondria, supporting nucleotide metabolism and immune responses. |
| GALNT2 | 0.7372 | 1.95E-05 | O-glycan biosynthesis | Initiates mucin-type O-glycosylation by transferring GalNAc to serine/threonine residues of proteins. |
| CSGALNACT2 | 1.1960 | 3.22E-04 | Glycosaminoglycan biosynthesis | Adds N-acetylgalactosamine in chondroitin sulphate biosynthesis, important in extracellular matrix formation. |

| | | | | |
|---|---|---|---|---|
| UGCG | 1.1699 | 6.49E-04 | Lactosylceramide biosynthesis | Catalyses the transfer of glucose to ceramide, forming glucosylceramide in glycosphingolipid biosynthesis. |
| B3GALNT1 | 1.7846 | 2.05E-04 | Glycosphingolipid biosynthesis | Transfers N-acetylgalactosamine to glycolipids during complex ganglioside biosynthesis. |
| MAN1A1 | 0.6051 | 2.71E-02 | N-glycan precursor trimming | Trims mannose residues from N-linked oligosaccharides in the Golgi apparatus during protein maturation. |
| MAN2A2 | 1.0553 | 1.32E-03 | N-glycan precursor trimming | Involved in the maturation of N-glycans by removing mannose residues from glycoproteins. |
| AGPAT2 | 0.5912 | 7.27E-03 | Triacylglycerol biosynthesis | Converts lysophosphatidic acid to phosphatidic acid, a key step in triglyceride and phospholipid synthesis. |
| CHKA | 1.6124 | 3.44E-06 | Phosphatidylcholine (PC) biosynthesis, Phosphatidylethanolamine (PE) biosynthesis | Catalyses the phosphorylation of choline to phosphocholine in the Kennedy pathway for phosphatidylcholine biosynthesis. |
| SPTLC2 | 1.0501 | 1.48E-05 | Ceramide biosynthesis, Sphingosine biosynthesis | Catalyses the initial step in sphingolipid biosynthesis by condensing serine with palmitoyl-CoA. |
| IDI1 | 0.7036 | 1.61E-03 | C5 isoprenoid biosynthesis, mevalonate pathway | Isomerises isopentenyl pyrophosphate to dimethylallyl pyrophosphate in the cholesterol biosynthetic pathway. |
| CEL | 1.2848 | 4.66E-03 | Acylglycerol degradation | Breaks down dietary fat and cholesterol esters in the digestive tract. |
| SPHK1 | 0.7244 | 2.47E-02 | Sphingosine degradation | Phosphorylates sphingosine to sphingosine-1-phosphate, a lipid signalling molecule involved in inflammation and cell survival. |
| EBP | 0.7447 | 1.48E-02 | Cholesterol biosynthesis | Encodes a sterol isomerase involved in the final steps of cholesterol biosynthesis. |
| HSD17B4 | 0.8070 | 4.87E-03 | Bile acid biosynthesis | Functions in peroxisomal β-oxidation and steroid metabolism by oxidising 17β-hydroxysteroids. |
| CYP19A1 | 5.6456 | 5.43E-06 | C19/C18-Steroid hormone biosynthesis | Encodes aromatase, which converts androgens into oestrogens. |
| GCLC | 1.4025 | 1.11E-04 | Glutathione biosynthesis | Catalyses the first and rate-limiting step in glutathione synthesis, essential for redox homeostasis. |
| COQ2 | 0.6041 | 7.00E-03 | Ubiquinone biosynthesis | Catalyses a step in the biosynthesis of coenzyme Q10 (ubiquinone), important in mitochondrial respiration. |
| PLCD3 | 1.3502 | 1.27E-02 | Inositol phosphate metabolism | Hydrolyses phosphatidylinositol 4,5-bisphosphate into diacylglycerol and IP3, mediating intracellular signalling. |

| | | | | |
|---|---|---|---|---|
| MAOA | 4.832<br>5 | 5.14E-<br>15 | GABA biosynthesis | Degrades monoamine neurotransmitters like dopamine, serotonin, and norepinephrine via oxidative deamination. |
| HK1 | 1.595<br>0 | 4.15E-<br>04 | Glycolysis, Nucleotide sugar biosynthesis | Phosphorylates glucose to glucose-6-phosphate in the first step of glycolysis. |
| GALM | 0.762<br>4 | 2.85E-<br>02 | Galactose degradation, Leloir pathway | Converts β-D-galactose to α-D-galactose, facilitating its entry into the Leloir pathway. |
| PCCB | 0.665<br>0 | 1.00E-<br>02 | Propanoyl-CoA metabolism | Catalyses the conversion of propionyl-CoA to methylmalonyl-CoA in the catabolism of odd-chain fatty acids. |
| GAPDH | 0.898<br>4 | 6.79E-<br>06 | Glycolysis, Gluconeogenesis | Catalyses the sixth step in glycolysis and also functions in nuclear transcription and apoptosis. |
| PGD | 0.814<br>9 | 1.41E-<br>02 | Pentose phosphate pathway | Catalyses an oxidative step in the pentose phosphate pathway, generating NADPH. |
| IDH2 | 1.010<br>6 | 6.89E-<br>05 | Citrate cycle (TCA cycle, Krebs cycle) | Mitochondrial isoform converting isocitrate to α-ketoglutarate while producing NADPH. |
| PSAT1 | 1.353<br>6 | 3.30E-<br>04 | Serine biosynthesis | Catalyses the conversion of 3-phosphohydroxypyruvate to phosphoserine in the serine synthesis pathway. |
| ARG2 | 1.606<br>6 | 1.36E-<br>02 | Urea cycle, Polyamine biosynthesis | Converts arginine into ornithine and urea in the mitochondria, functioning in extrahepatic tissues. |
| GSTZ1 | 0.613<br>9 | 1.13E-<br>02 | Tyrosine degradation | Detoxifies electrophilic compounds through glutathione conjugation, including in tyrosine degradation. |
| NME4 | 0.866<br>7 | 2.63E-<br>02 | Purine metabolism, Pyrimidine ribonucleotide biosynthesis | Mitochondrial nucleoside diphosphate kinase involved in mitochondrial DNA replication and repair. |
| CTPS1 | 0.655<br>2 | 2.38E-<br>02 | Pyrimidine ribonucleotide biosynthesis | Catalyses the rate-limiting step in the de novo synthesis of CTP from UTP, essential for DNA/RNA synthesis. |
| GCNT1 | 1.039<br>7 | 1.82E-<br>05 | O-glycan biosynthesis | Catalyses the branching of O-glycans by transferring N-acetylglucosamine, influencing cell adhesion and signalling. |
| B4GALT3 | 0.945<br>3 | 3.49E-<br>02 | N-glycan precursor trimming | Adds galactose to N-acetylglucosamine in glycoproteins and glycolipids, important in cell-cell communication. |
| CHPT1 | 1.232<br>2 | 1.96E-<br>03 | Phosphatidylcholine (PC) biosynthesis | Involved in phosphatidylcholine biosynthesis by transferring phosphocholine to diacylglycerol. |

| | | | | |
|---|---|---|---|---|
| ACAT1 | 0.912 6 | 1.60E-03 | C5 isoprenoid biosynthesis, mevalonate pathway | Converts acetyl-CoA to acetoacetyl-CoA, playing a role in ketone body metabolism and cholesterol synthesis. |
| PNPLA2 | 0.795 6 | 1.75E-02 | Acylglycerol degradation | Encodes adipose triglyceride lipase, responsible for hydrolysing stored triglycerides into free fatty acids. |
| CYP51A1 | 1.036 4 | 2.21E-04 | Cholesterol biosynthesis | Lanosterol 14α-demethylase involved in the conversion of lanosterol to cholesterol. |
| HSD3B7 | 1.946 3 | 4.35E-06 | Bile acid biosynthesis | Catalyses the first oxidative step in bile acid biosynthesis from cholesterol. |
| COQ3 | 0.993 1 | 1.60E-02 | Ubiquinone biosynthesis | O-methyltransferase is involved in coenzyme Q10 biosynthesis, essential for mitochondrial electron transport. |
| MAOB | 1.564 9 | 2.51E-02 | GABA biosynthesis | Oxidises neurotransmitters like dopamine and serotonin, playing a role in mood and neurodegeneration. |
| HK3 | 1.622 9 | 3.38E-05 | Glycolysis, Nucleotide sugar biosynthesis | A hexokinase isoform that phosphorylates glucose in the initial step of glycolysis, especially in immune cells. |
| GALE | 0.659 0 | 2.47E-02 | Galactose degradation (Leloir pathway) | Interconverts UDP-galactose and UDP-glucose, critical for galactose metabolism. |
| PKM | 0.649 9 | 1.14E-05 | Glycolysis | Pyruvate kinase isozyme catalyses the final step of glycolysis, generating ATP and pyruvate. |
| ASS1 | 3.586 6 | 1.82E-02 | Urea cycle | Synthesises argininosuccinate from citrulline and aspartate in the urea cycle. |
| HPD | 2.008 6 | 1.49E-02 | Tyrosine degradation | Converts 4-hydroxyphenylpyruvate to homogentisate in the catabolic pathway of tyrosine. |
| GMPS | 0.587 9 | 2.42E-02 | Purine metabolism | Converts xanthosine monophosphate to guanosine monophosphate in purine biosynthesis. |
| DHCR7 | 0.930 5 | 7.73E-03 | Cholesterol biosynthesis | Catalyses the conversion of 7-dehydrocholesterol to cholesterol, the final step in cholesterol synthesis. |
| PI4K2B | 0.966 4 | 1.54E-05 | Inositol phosphate metabolism | Produces phosphatidylinositol 4-phosphate, important for vesicle trafficking and membrane identity. |
| GALNT10 | 0.589 0 | 2.85E-02 | O-glycan biosynthesis | Initiates O-linked glycosylation by transferring GalNAc to serine/threonine residues of proteins. |
| DHCR24 | 2.494 6 | 4.02E-09 | Cholesterol biosynthesis | Reduces desmosterol to cholesterol and is involved in anti-apoptotic signalling. |

| | | | | |
|---|---|---|---|---|
| ITPKC | 1.0837 | 6.29E-04 | Inositol phosphate metabolism | Regulates calcium signalling by phosphorylating inositol 1,4,5-trisphosphate. |
| GALNT14 | 2.2820 | 3.15E-04 | O-glycan biosynthesis | Initiates mucin-type O-glycosylation and is associated with apoptotic regulation. |
| MSMO1 | 0.7460 | 1.08E-04 | Cholesterol biosynthesis | Involved in cholesterol biosynthesis by demethylating sterol intermediates. |
| PIP5K1B | 0.9984 | 1.54E-02 | Inositol phosphate metabolism | Catalyses formation of phosphatidylinositol 4,5-bisphosphate, regulating cytoskeletal dynamics and signalling. |
| PFKP | 0.6030 | 8.01E-04 | Glycolysis | A key rate-limiting enzyme in glycolysis converting fructose-6-phosphate to fructose-1,6-bisphosphate. |
| SQLE | 1.3534 | 6.83E-04 | Cholesterol biosynthesis | Squalene monooxygenase, catalysing a rate-limiting step in cholesterol biosynthesis. |
| TM7SF2 | 0.7829 | 1.09E-02 | Cholesterol biosynthesis | Involved in the final steps of cholesterol biosynthesis, particularly sterol reduction. |
| PIGX | -0.8741 | 2.83E-05 | GPI-anchor biosynthesis | Participates in early steps of glycosylphosphatidylinositol (GPI) anchor biosynthesis. |
| B3GALT2 | -1.6295 | 1.02E-03 | Glycosphingolipid biosynthesis | Transfers galactose to N-acetylglucosamine in glycosphingolipid biosynthesis. |
| IDS | -0.7798 | 7.18E-04 | Heparan sulphate degradation | Encodes iduronate-2-sulfatase, which degrades dermatan and heparan sulphate in lysosomes. |
| PPCDC | -0.8863 | 4.41E-05 | Coenzyme A biosynthesis | Catalyses the decarboxylation step in the biosynthesis of coenzyme A from pantothenate. |
| IPMK | -0.6055 | 3.83E-02 | Inositol phosphate metabolism | Phosphorylates inositol phosphates and also regulates transcription and energy metabolism. |
| MAN1C1 | -1.0267 | 1.90E-03 | N-glycan precursor trimming | Removes mannose residues during N-glycan maturation in the ER-Golgi pathway. |
| PLCB1 | -0.9966 | 1.49E-03 | Inositol phosphate metabolism | Cleaves phosphatidylinositol 4,5-bisphosphate to generate DAG and IP3 for calcium signalling. |
| GCNT4 | -1.0655 | 2.79E-03 | O-glycan biosynthesis | Adds N-acetylglucosamine to O-glycans, contributing to core 2 branched structures on glycoproteins. |

**Table S4: Comparison of predictive model performance using serum metabolites from adults with HLH vs. sepsis and rheumatic disease controls.** Performance statistics for 6 predictive models based on serum metabolites, demographic and clinical features (where available) for the HLH vs. SLE and HLH vs. RA. Additional models were generated to explore the influence of sex (SLE) and age / clinical features (RA) on features selected by the models. The models used were Lasso LR with and without interactions (I), NN, RF, SVM, and XGBoost. The sensitivity represents the true positive rate (HLH) in contrast to specificity, which is the true negative rate (rheumatic disease controls: SLE, RA). Statistics were rounded to four decimal places. Refer to Table 2, Figure 4 and Figures S3 and S4.

| Comparison | Model | Accuracy | Sensitivity | Specificity | AUC-ROC | MCC |
|---|---|---|---|---|---|---|
| **HLH vs. Sepsis** (excluding treatment and age**)** | LR | 0.7315 | 0.6667 | 0.8039 | 0.8000 | 0.4728 |
| | LR+I | 0.7407 | 0.7018 | 0.7843 | 0.8100 | 0.4861 |
| | NN | 0.7685 | 0.7719 | 0.7647 | 0.8300 | 0.5362 |
| | RF | 0.7130 | 0.7368 | 0.6863 | 0.7500 | 0.4236 |
| | SVM | 0.6111 | 0.5439 | 0.6863 | 0.6300 | 0.2317 |
| | XGBoost | 0.7407 | 0.7018 | 0.7843 | 0.8100 | 0.4861 |
| **HLH vs. Sepsis** (metabolites only) | LR | 0.6574 | 0.6491 | 0.6667 | 0.7300 | 0.3153 |
| | LR+I | 0.6944 | 0.6667 | 0.7255 | 0.8100 | 0.3918 |
| | NN | 0.7315 | 0.7719 | 0.6863 | 0.8200 | 0.4603 |
| | RF | 0.6852 | 0.7193 | 0.6471 | 0.7400 | 0.3674 |
| | SVM | 0.5926 | 0.5614 | 0.6275 | 0.6100 | 0.1889 |
| | XGBoost | 0.7037 | 0.7368 | 0.6667 | 0.7800 | 0.4046 |
| **HLH vs. Sepsis** (clinical lab markers only*) | LR | 0.7037 | 0.8421 | 0.5490 | 0.7900 | 0.4114 |
| | LR+I | 0.7130 | 0.7544 | 0.6667 | 0.7900 | 0.4230 |
| | NN | 0.7500 | 0.8421 | 0.6471 | 0.7700 | 0.5009 |
| | RF | 0.7315 | 0.7018 | 0.7647 | 0.7700 | 0.4661 |
| | SVM | 0.7315 | 0.8246 | 0.6275 | 0.7800 | 0.4629 |
| | XGBoost | 0.7222 | 0.7368 | 0.7059 | 0.7400 | 0.4427 |
| **HLH vs. SLE** (excluding Sex) | LR | 0.9048 | 0.8596 | 0.9279 | 0.9652 | 0.7876 |
| | LR+I | 0.8810 | 0.8070 | 0.9189 | 0.9567 | 0.7324 |
| | NN | 0.9107 | 0.8947 | 0.9189 | 0.9458 | 0.8040 |
| | RF | 0.9107 | 0.9474 | 0.8919 | 0.9474 | 0.8136 |
| | SVM | 0.8929 | 0.8421 | 0.9189 | 0.9469 | 0.7610 |
| | XGBoost | 0.8869 | 0.8772 | 0.8919 | 0.8919 | 0.7546 |
| **HLH vs. RA** (excluding lab markers and age) | LR | 0.9385 | 0.9474 | 0.9348 | 0.9349 | 0.8580 |
| | LR+I | 0.9795 | 0.9474 | 0.9928 | 0.9967 | 0.9502 |
| | NN | 0.9846 | 0.9474 | 1.0000 | 0.9950 | 0.9629 |
| | RF | 0.9692 | 0.9474 | 0.9783 | 0.9898 | 0.9256 |
| | SVM | 0.9744 | 0.9474 | 0.9855 | 0.9933 | 0.9378 |
| | XGBoost | 0.9179 | 0.9123 | 0.9203 | 0.9836 | 0.8097 |

**Table S5: Pathways identified by metabolite set enrichment analysis (MSEA) analysis that were shared between the HLH vs. sepsis, SLE and RA.** Over-representation analysis was performed on metabolomic signatures associated with HLH vs sepsis, SLE and RA identified by at least one ML model, multiple Mann-Whitney U tests and univariate logistic regression using the Relational Database of Metabolomics Pathways tool within MetaboAnalyst 6.0 (13). Metabolic pathways with significance of FDR<0.05 were included**.** Table displays the top 25 pathways identified in the HLH vs sepsis comparison. These pathways were also significantly over-represented in the HLH vs SLE and HLH vs RA comparisons. Refer to Figure 4, Figures S3 & S4.

| **Pathway** | **P value (FDR)** | | |
|---|---|---|---|
| | **HLH vs Sepsis** | **HLH vs SLE** | **HLH vs RA** |
| Familial hyperlipidemia type 2 | 8.32E-14 | 4.16E-14 | 1.82E-11 |
| Statin inhibition of cholesterol production | 9.27E-14 | 7.10E-12 | 5.93E-08 |
| Familial hyperlipidemia type 4 | 1.01E-13 | 1.01E-13 | 2.60E-09 |
| Familial hyperlipidemia type 3 | 8.35E-13 | 8.35E-13 | 2.39E-10 |
| Transport of small molecules | 1.25E-12 | 4.06E-15 | 2.84E-11 |
| Familial hyperlipidemia type 5 | 6.22E-12 | 7.10E-12 | 1.43E-09 |
| Familial hyperlipidemia type 1 | 6.22E-12 | 7.10E-12 | 1.43E-09 |
| Lipid particles composition | 6.22E-12 | 1.54E-09 | 2.80E-05 |
| SLC-mediated transmembrane transport | 1.76E-07 | 1.02E-11 | 1.89E-10 |
| LDL clearance | 2.06E-07 | 1.46E-05 | 2.84E-02 |
| Plasma lipoprotein clearance | 2.06E-07 | 1.46E-05 | 2.84E-02 |
| VLDL assembly | 2.09E-07 | 6.57E-05 | 6.81E-03 |
| Chylomicron assembly | 2.09E-07 | 6.57E-05 | 6.81E-03 |
| HDL clearance | 2.09E-07 | 6.57E-05 | 6.81E-03 |
| Chylomicron clearance | 2.09E-07 | 6.57E-05 | 6.81E-03 |
| LDL remodeling | 2.09E-07 | 6.57E-05 | 6.81E-03 |
| VLDL clearance | 2.09E-07 | 6.57E-05 | 6.81E-03 |
| Disorders of transmembrane transporters | 4.97E-07 | 2.85E-11 | 3.52E-11 |
| Metabolic pathway of LDL, HDL, TG, including diseases | 8.83E-07 | 8.83E-07 | 8.79E-07 |
| SREBP signaling | 8.83E-07 | 8.83E-07 | 1.09E-04 |
| Cholestasis | 1.08E-06 | 6.57E-05 | 8.29E-05 |
| Transport of bile salts, organic acids, metal ions, amine compounds | 1.24E-06 | 5.43E-11 | 5.93E-08 |
| SLC transporter disorders | 1.79E-06 | 8.91E-11 | 9.57E-11 |
| Metabolic reprogramming in pancreatic cancer | 2.01E-06 | 2.10E-06 | 1.05E-03 |
| Plasma lipoprotein remodeling | 2.03E-06 | 1.08E-04 | 6.18E-02 |

**Table S6: List of metabolites identified by either ML, univariate logistic regression or differential metabolite abundance analysis that can stratify HLH from controls.** Metabolites in bold have AUCs greater than 0.70 across all comparisons. Only metabolite concentrations included. Ranked according to AUC values for the HLH vs Sepsis comparison. PS-M, Post-surgery malignancy. See Table S2 for metabolite abbreviations. *metabolites selected for more detailed analysis in Figure 5 and Figure S5).

| Metabolite | HLH vs. Sepsis | HLH vs. SLE | HLH vs. RA | HLH vs. PS-M |
|---|---|---|---|---|
| **IDL-P*** | 0.7957 | 0.7525 | 0.8072 | 0.8289 |
| **S-LDL-CE*** | 0.7910 | 0.7013 | 0.7597 | 0.8026 |
| **XS-VLDL-C*** | 0.7802 | 0.8649 | 0.9485 | 0.9649 |
| **XS-VLDL-P*** | 0.7786 | 0.9495 | 0.9943 | 1.0000 |
| **XS-VLDL-L*** | 0.7755 | 0.9516 | 0.9920 | 1.0000 |
| **S-LDL-L** | 0.7570 | 0.7568 | 0.8272 | 0.8421 |
| **XS-VLDL-CE** | 0.7523 | 0.7582 | 0.8587 | 0.8904 |
| **XS-VLDL-FC** | 0.7523 | 0.9417 | 0.9908 | 1.0000 |
| **LDL-P** | 0.7492 | 0.7610 | 0.8587 | 0.8509 |
| **LDL-TG*** | 0.7461 | 0.9858 | 0.9966 | 1.0000 |
| **S-VLDL-CE** | 0.7446 | 0.9111 | 0.9611 | 0.9649 |
| **ApoB*** | 0.7430 | 0.8037 | 0.8959 | 0.8684 |
| **M-LDL-TG** | 0.7430 | 0.9815 | 0.9966 | 1.0000 |
| **M-LDL-P** | 0.7400 | 0.7048 | 0.8089 | 0.8114 |
| **L-LDL-P** | 0.7368 | 0.7866 | 0.8930 | 0.8596 |
| **XS-VLDL-PL** | 0.7353 | 0.9644 | 0.9926 | 1.0000 |
| **L-LDL-TG** | 0.7322 | 0.9858 | 0.9977 | 1.0000 |
| **S-VLDL-C** | 0.7307 | 0.8940 | 0.9554 | 0.9649 |
| **L-HDL-TG** | 0.7307 | 0.8869 | 0.9199 | 1.0000 |
| **XL-HDL-TG** | 0.7291 | 0.9559 | 0.9903 | 1.0000 |
| **XS-VLDL-TG** | 0.7260 | 0.9844 | 0.9931 | 1.0000 |
| S-HDL-L | 0.7245 | 0.6686 | 0.8839 | 0.7807 |
| **SFA*** | 0.7214 | 0.8706 | 0.9136 | 0.9561 |
| **IDL-TG*** | 0.7214 | 0.9858 | 0.9966 | 1.0000 |
| **S-LDL-P*** | 0.7198 | 0.7511 | 0.8633 | 0.8333 |
| **S-HDL-P** | 0.7121 | 0.7688 | 0.9125 | 0.8904 |
| **HDL-TG*** | 0.7105 | 0.9403 | 0.9640 | 0.9956 |
| **VLDL-CE** | 0.7043 | 0.8407 | 0.9416 | 0.9474 |
| M-HDL-TG | 0.6981 | 0.9168 | 0.9394 | 0.9825 |
| S-VLDL-FC | 0.6950 | 0.8478 | 0.9359 | 0.9386 |
| XL-HDL-P | 0.6625 | 0.7496 | 0.6779 | 0.8026 |
| Lactate | 0.6440 | 0.6444 | 0.7866 | 0.8640 |
| Phe | 0.6217 | 0.9020 | 0.9285 | 0.9737 |
| XL-VLDL-TG | 0.6192 | 0.8492 | 0.9325 | 0.7982 |
| Unsaturation | 0.5836 | 0.6935 | 0.9703 | 0.9693 |

**Table S7: ROC analysis was performed to assess the ability of ApoB, ApoA1 and the ApoB/ApoA1 ratio to discriminate HLH (n=26) from sepsis (n=22) using Youden index.**

| Parameter | Cut-off | Youden J statistic | Accuracy | Sensitivity | Specificity | AUC |
|---|---|---|---|---|---|---|
| **ApoB/ApoA1** | 0.2757 | 0.4266 | 0.7083 | 0.6538 | 0.7727 | 0.7867 |
| **ApoB** | 554350 | 0.4196 | 0.7083 | 0.6923 | 0.7273 | 0.7465 |
| **ApoA1** | 2604200 | -0.021 | 0.5 | 0.6154 | 0.3636 | 0.5962 |

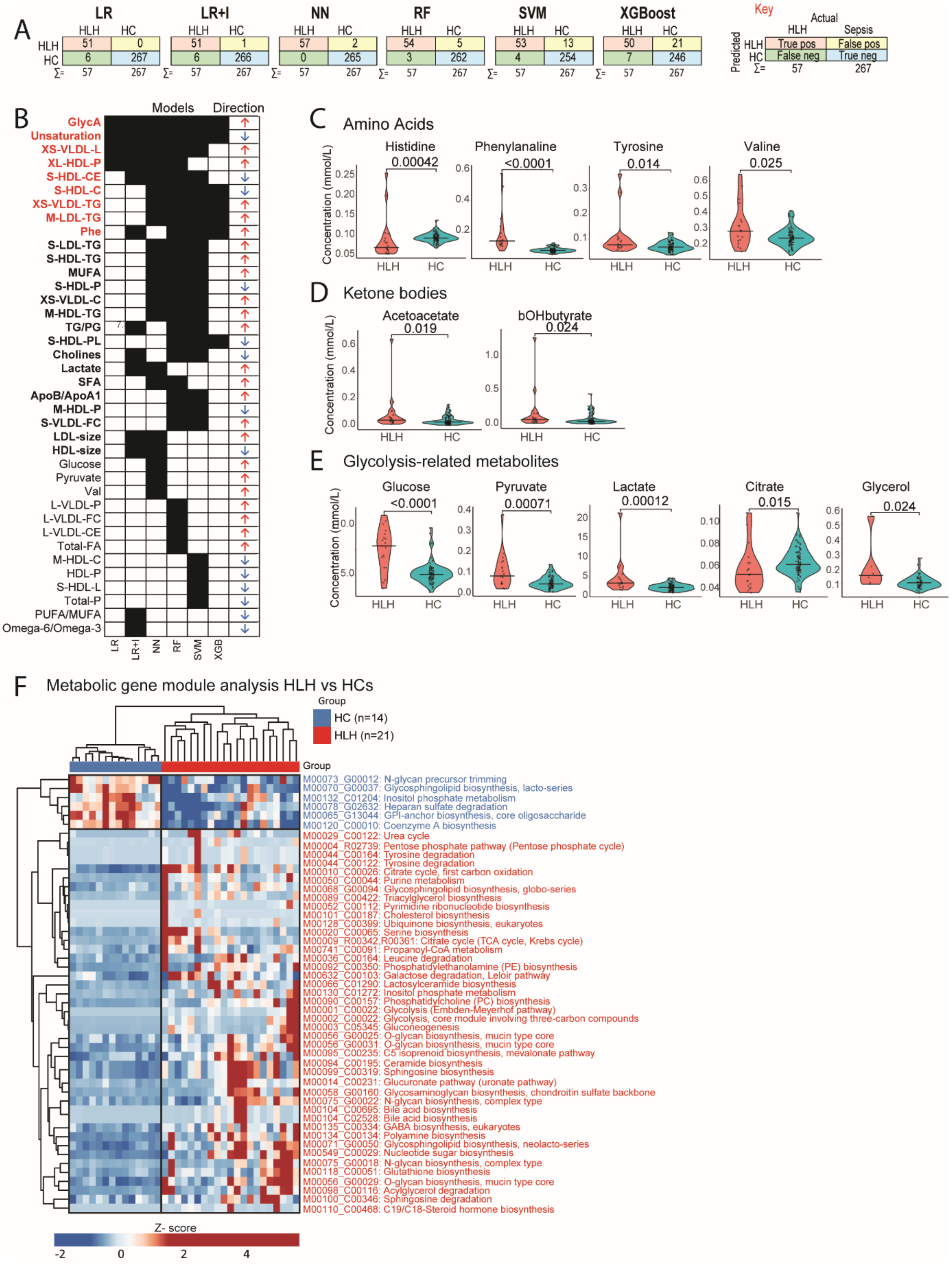


**Figure S1: Adults hospitalised with HLH are characterised by changes in metabolism.** Sera from adults hospitalised with HLH (n=19) and healthy controls (n=89) were analysed using an NMR metabolomic platform. (**A**) Metabolomic signatures associated with HLH vs. HCs were determined using multiple machine learning (ML) models: logistic regression (LR) with/without interactions (I), neural network (NN), random forest (RF), support vector machine (SVM) and eXtreme Gradient Boosting (XGB). Confusion matrices showing the number of correct (red and blue squares) and incorrect (yellow and green squares) classifications for each model. **(B)** Comparison of metabolites selected by each ML model (black squares). Metabolites selected by ≥ 4 models (red bold); metabolites selected by 2-3 models (black bold). Arrows represent whether the metabolite/clinical feature is elevated (↑) or lower (↓) in HLH vs HCs. **(C-E)** Violin plots showing the concentration or ratio of metabolites in serum from HLH patients (red) and HCs (turquoise). **(C)**; Amino acids; **(D)** Ketone bodies **(E)** Glycolysis-related metabolites. Mann-Whitney U tests, with p-value, each data point and median shown. See Figure 2A-E. **(F)** Whole blood RNA-sequencing was performed on matched samples (HLH, n=21; and HCs, n=14). Metabolic gene module analysis identified 75 differentially expressed genes were associated with 49 metabolic modules (subnetworks or processes within a metabolic pathway) in HLH vs. HCs. Heatmap of 49 dysregulated metabolic modules between HLH patients and HCs. The 43 upregulated and downregulated modules in HLH patients compared to HCs are highlighted in red and blue, respectively (see Figure 3, Table S3).

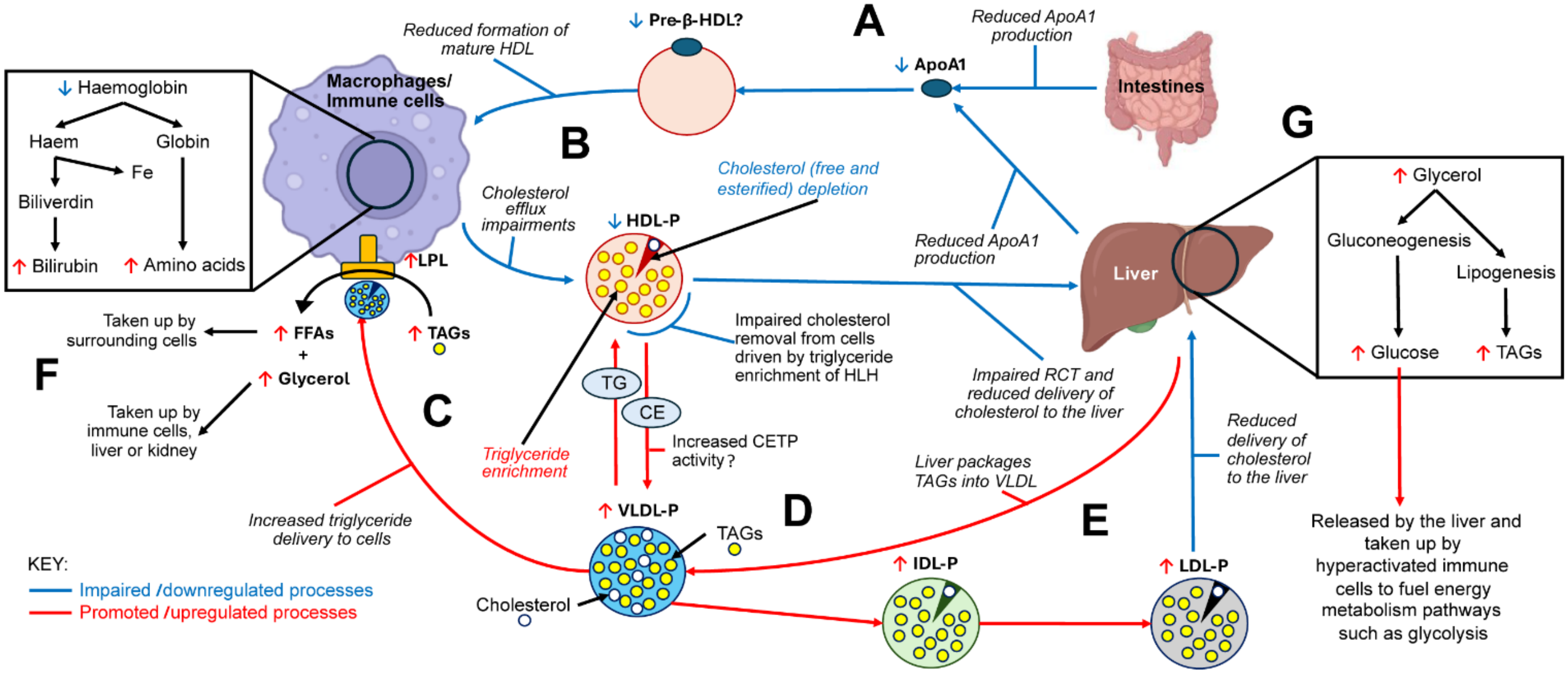


**Figure S2: Proposed mechanism based on metabolomic and transcriptomic data for lipoprotein remodelling and impaired cholesterol efflux in adults with HLH**. **(A)** Low levels of ApoA1 may reduce the production of pre-β-HDL/nascent HDL. **(B)** Together with lipoprotein remodelling toward triglyceride enrichment and cholesterol-phospholipid depletion, this can impair cholesterol efflux from cells such as macrophages and disrupt reverse cholesterol transport (RCT), the process by which excess cholesterol is removed from peripheral tissues and returned to the liver for excretion via bile and faeces. As a result, cholesterol can potentially accumulate within cells. Effective efflux requires lipid acceptors such as HDL. **(C)** Elevated serum levels of ApoB-containing lipoproteins (VLDL-P, IDL-P, LDL-P) further contribute to metabolic dysregulation. VLDL particles deliver excess triglycerides (TAGs/TG) to cells, where they are hydrolysed into triglycerides and free fatty acids (FFAs). Triglycerides are subsequently taken up by immune cells, the liver, or the kidney, while FFAs are absorbed by surrounding cells, including macrophages. **(D)** Transfer of triglycerides from VLDL to HDL particles and movement of cholesterol esters (CE)

from HDL to VLDL. **(E)** LDL-P, which is depleted of cholesterol, may lead to reduced delivery of cholesterol to the liver for clearance. **(F)** Within macrophages, haemoglobin is normally degraded into haem and globin. Globin is broken down into amino acids, while haem is converted into biliverdin and then bilirubin **(G)** Glycerol in the liver can enter gluconeogenesis to generate glucose for immune cell energy demands or be channelled into lipogenesis for the synthesis of new triglycerides, which are packaged into VLDL particles. ↑ = elevated within the serum of hosp-HLH patients, ↓ = lower in the serum of hosp-HLH patients. Created using biorender

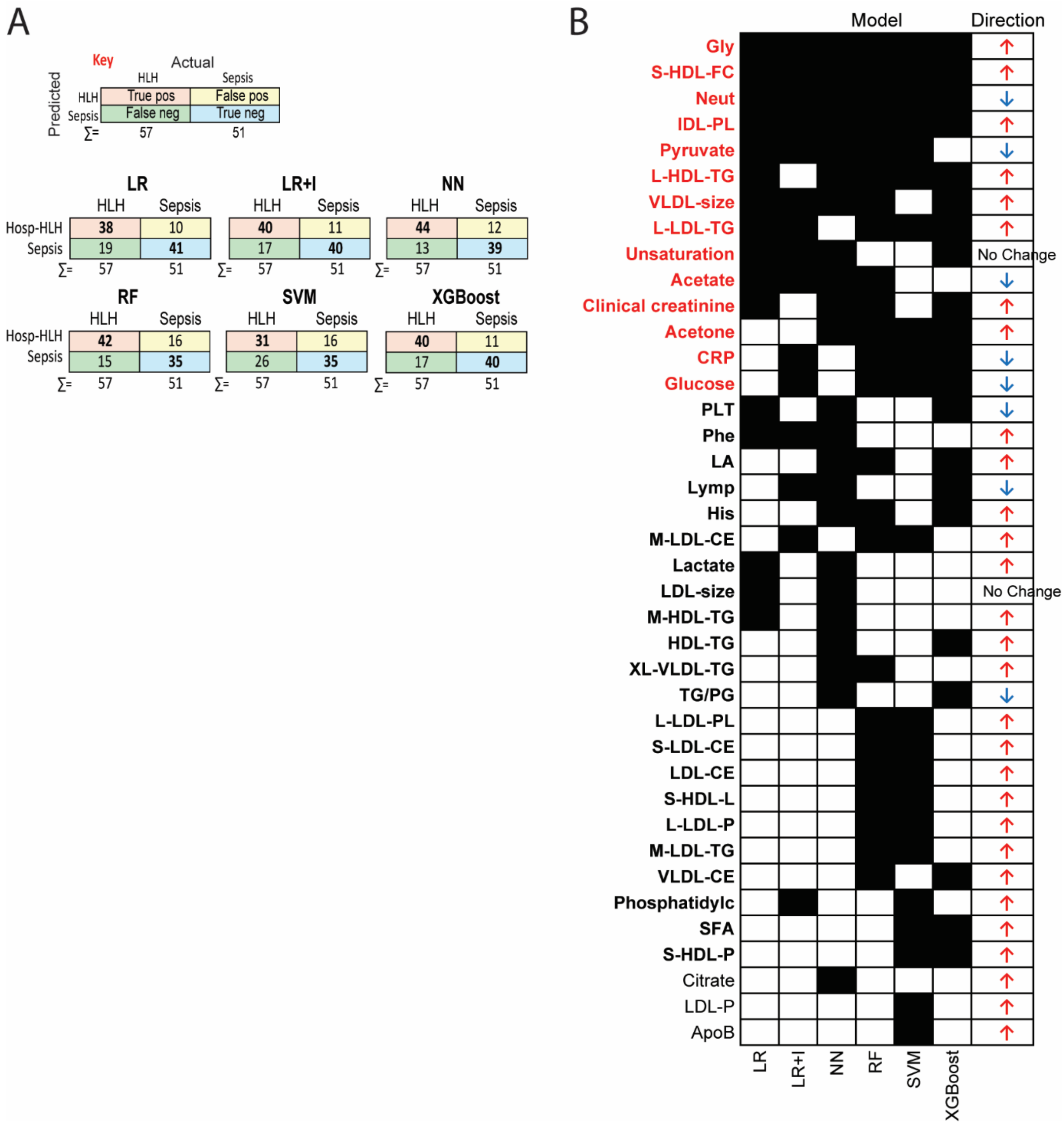


**Figure S3**: **Serum metabolomics can stratify adults hospitalised with HLH from those with sepsis (excluding age and treatment e.g. systemic steroid).** NMR serum metabolomics analysis in patients with HLH (n=19) and sepsis (n=17). Metabolomic signatures associated with HLH vs. sepsis were determined using machine learning. (**A**) Confusion matrices showing the number of correct (red and blue squares) and incorrect (yellow and green squares) classifications for each model. The sum (Σ) of each row and column is given. The models were: logistic regression (LR) with/without interactions (I), neural network (NN), random forest (RF), support vector machine (SVM) and eXtreme Gradient Boosting (XGBoost). As SVM performed poorly at stratifying both patient groups, it was excluded from downstream analyses. (**B**) Comparison of metabolites selected by each machine learning model (black squares). Metabolites selected by ≥ 4 models highlighted in red bold; metabolites selected by 2-3 models in black bold (see Table S4, Figure 4A-E).

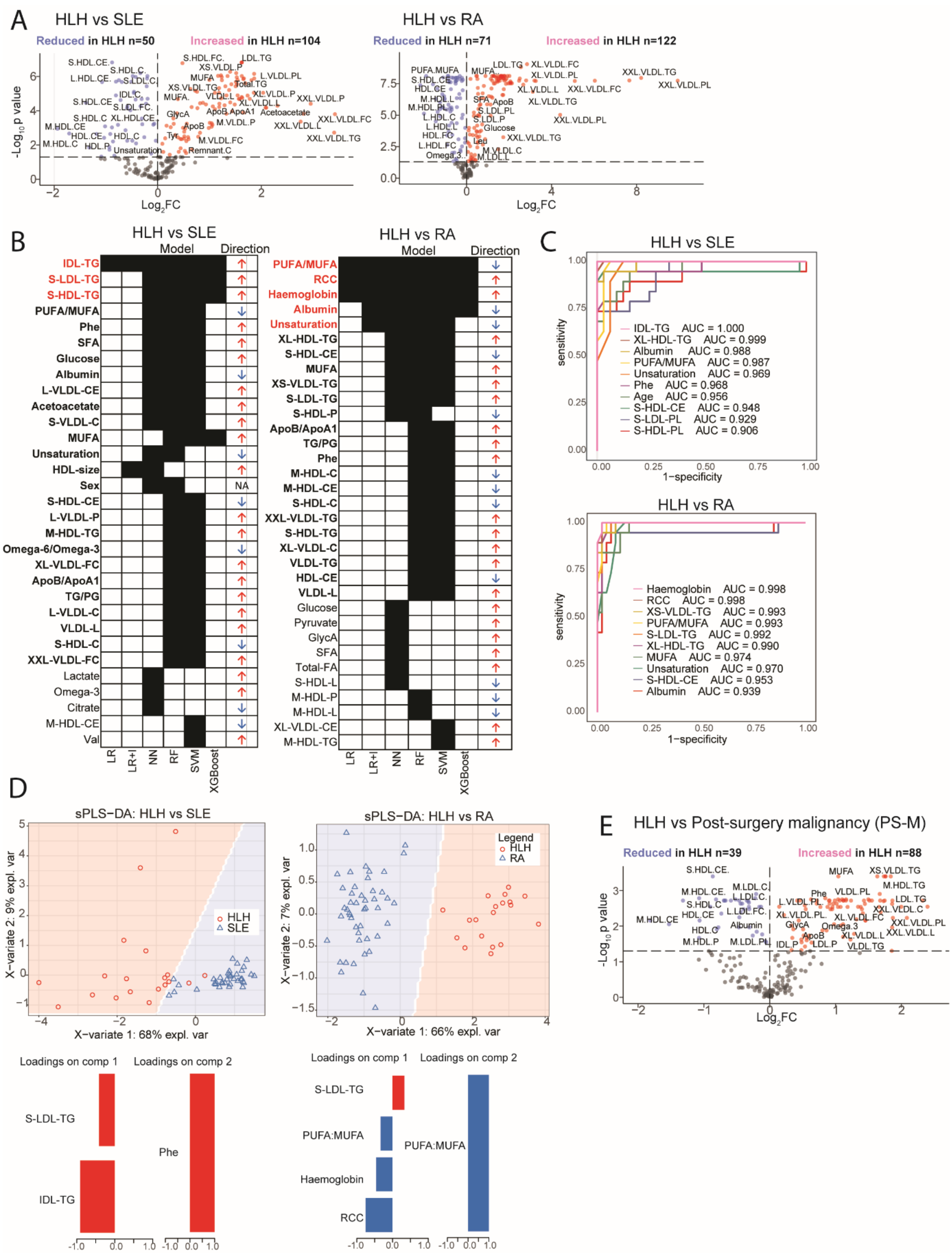


**Figure E4: Serum metabolomics can stratify adults hospitalised with HLH from those with systemic lupus erythematosus (SLE) and rheumatoid arthritis (RA). (A)** Volcano plots showing differentially abundant metabolites in HLH (n=19) vs. SLE (n=37)

and HLH (n=19) vs. RA (n=46). **(B-D)** Metabolomic signatures associated with HLH (n = 19) vs SLE (n=37) and HLH vs RA (n=46) were determined using machine learning (ML) models: logistic regression (LR) with/without interactions (I), neural network (NN), random forest (RF), support vector machine (SVM) and eXtreme Gradient Boosting (XGBoost). **(B)** Comparison of features selected by each ML model (black squares). Metabolites selected by ≥4 models in red bold; metabolites selected by 2-3 models in black bold. Arrows represent whether feature is elevated (↑) or lower (↓) in HLH vs SLE (left) and RA (right). (**C**) Area under the curve-Receiver operator characteristic (AUC-ROC) of top 10 metabolites/clinical features in HLH vs SLE (top) and HLH vs RA (bottom). (**D**) sPLS-DA plot (sparse partial least squares-discriminant analysis) to validate metabolomic signatures i.e. top 9 metabolites (ex. age) identified by ML in HLH vs SLE (left) and HLH vs RA (right). Features in components 1 and 2 driving the separation are shown. **(E)**) Volcano plots showing differentially abundant metabolites in HLH (n=19) vs. post-surgery malignancy, (PS-M) (n=6). See Table S2 for metabolite abbreviations.

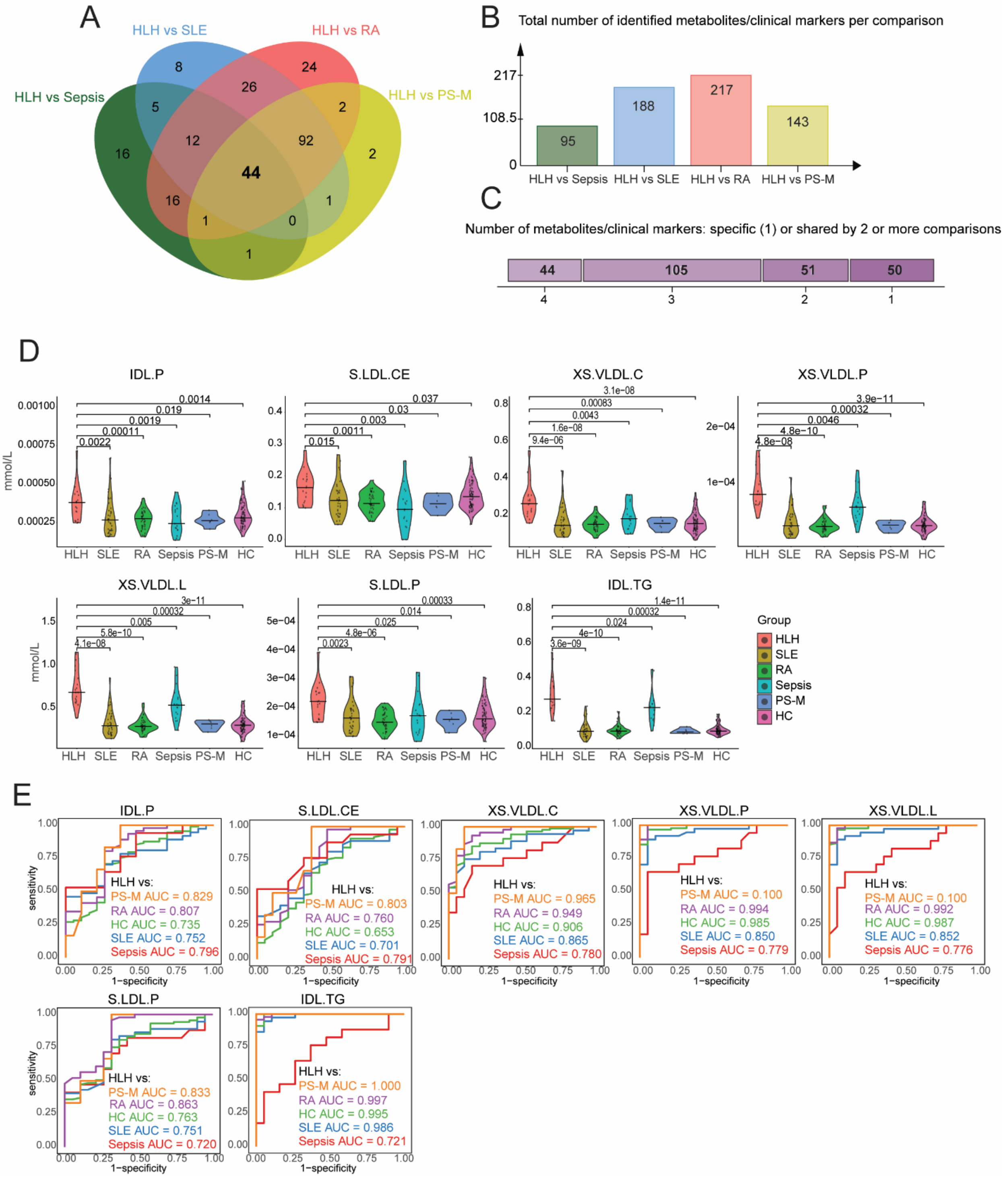


**Figure E5: Identification of shared and metabolites across HLH comparisons with disease controls.** (**A**) Venn diagram showing the number of shared metabolites across the following comparisons: HLH vs sepsis (dark green), HLH vs SLE (blue), HLH vs RA (red) and HLH vs post-surgery malignancy (PS-M, yellow). (**B**) Total number of metabolites and clinical markers identified in each comparison. (**C**) Number of metabolites and clinical markers that were either unique to a single comparison or shared across multiple comparisons (ranging from 2 to 6 comparisons). **(D)** Violin plots showing selected metabolomic biomarkers (from Table S6) that stratify HLH patients from all disease controls. P-values computed using Mann-Whitney U tests. Refer to Figure 5A. (**E**) Multi-ROC curves demonstrating how each metabolomic biomarker stratifies HLH from each disease control. Refer to Figure 5B.
